# Immune cell topography predicts response to PD-1 blockade in cutaneous T cell lymphoma

**DOI:** 10.1101/2020.12.06.20244913

**Authors:** Darci Phillips, Magdalena Matusiak, Belén Rivero Gutierrez, Salil S. Bhate, Graham L. Barlow, Sizun Jiang, Janos Demeter, Kimberly S. Smythe, Robert H. Pierce, Steven P. Fling, Nirasha Ramchurren, Martin A. Cheever, Yury Goltsev, Robert B. West, Michael S. Khodadoust, Youn H. Kim, Christian M. Schürch, Garry P. Nolan

## Abstract

Anti-PD-1 immunotherapies have transformed cancer treatment, yet the determinants of clinical response are largely unknown. We performed CODEX multiplexed tissue imaging and RNA sequencing on 70 tumor regions from 14 advanced cutaneous T cell lymphoma (CTCL) patients enrolled in a clinical trial of pembrolizumab therapy. Clinical response was not associated with the frequency of tumor-infiltrating T cell subsets, but rather with striking differences in the spatial organization and functional immune state of the tumor microenvironment (TME). After treatment, pembrolizumab responders had a localized enrichment of tumor and CD4^+^ T cells, which coincided with immune activation and cytotoxic PD-1^+^ CD4^+^ T cells. In contrast, non-responders had a localized enrichment of Tregs pre- and post-treatment, consistent with a persistently immunosuppressed TME and exhausted PD-1^+^ CD4^+^ T cells. Integrating these findings by computing the physical distances between PD-1^+^ CD4^+^ T cells, tumor cells, and Tregs revealed a spatial biomarker predictive of pembrolizumab response. Finally, the chemokine CXCL13 was upregulated in tumor cells in responders post-treatment, suggesting that chemoattraction of PD-1^+^ CD4^+^ T cells towards tumor cells facilitates a positive outcome. Together, these data show that T cell topography reflects the balance of effector and suppressive activity within the TME and predicts clinical response to PD-1 blockade in CTCL.

## INTRODUCTION

Cutaneous T cell lymphomas (CTCL) are a rare, heterogeneous group of T cell malignancies that primarily occur in the skin. The majority of CTCLs are comprised of mycosis fungoides and Sézary syndrome, which originate from mature, skin-tropic CD4^+^ T cells^1^. A third of patients present with advanced-stage disease and those meeting high risk criteria have a 5-year survival rate of 28%^2^. With the exception of hematopoietic stem cell transplantation, there are no curative therapies for advanced CTCL, and current treatments typically induce short-lived, partial disease control^3^. However, immune checkpoint inhibitors, such as antibodies against PD-1, restore T cell effector function at the tumor site^4,5^ and promote robust and durable responses in a number of advanced cancers^6-8^. In CTCL, PD-1 and its ligands can be simultaneously expressed on tumor cells, making this pathway an attractive therapeutic target for PD-1 blockade^9-12^.

A Cancer Immunotherapy Trials Network (CITN) multicenter phase II clinical trial of the anti-PD-1 immunotherapy, pembrolizumab, in heavily pre-treated patients with advanced relapsed/refractory CTCL was recently reported (NCT02243579)^13^. In this study, 38% of patients achieved a sustained clinical response, whereas 25% experienced rapid disease progression, likely because inhibition of PD-1 on tumor cells promoted cancer growth^14,15^. Such outcome discrepancies underscore a need for predictive biomarkers of PD-1 blockade that will allow patients to be stratified into probable responders and non-responders prior to initiating therapy.

To date, biomarker studies with immunohistochemistry (IHC), gene expression profiling, and mass cytometry (CyTOF), have not yet provided predictive indices of pembrolizumab responsiveness in CTCL^13^. Similarly, predictive biomarkers remain in-demand for other tumor types and immunotherapies^16,17^. As the immune system acts via coordinated cell-cell association, it is expected that spatial cellular attributes within the tumor microenvironment (TME) would be predictive of clinical outcomes. As such, organized cellular contexts would be productive whereas incorrectly arranged cellular contexts (i.e., absence or misplacement of certain cell-types) would prognosticate a negative outcome. Indeed, recent studies indicate that immune cells are not randomly distributed within the TME, but are purposefully organized into cellular neighborhoods and niches that facilitate anti- or pro-tumor functions^18,19^. This raises the question of how PD-1 blockade alters spatial cellular context, and in turn, how such changes in architecture relate to patient responses in CTCL.

We combined CO-Detection by indEXing (CODEX) multiplexed tissue imaging^19,20^ with gene expression profiling by RNA-seq to interrogate tumors from 14 patients with advanced CTCL sampled before and after pembrolizumab treatment. Then we computationally correlated spatial protein information at the single cell level with matched global gene expression data to generate insights into the drivers and resistors of PD-1 blockade. Our data revealed a localized enrichment of effector CD4^+^ T cells in responders versus inhibitory Tregs in non-responders. Such topographical differences coincided with variations in the functional immune state of the TME, T cell function, and tumor cell-specific chemokine expression. We integrated these findings into a single *SpatialScore*—a spatial biomarker predictive of pembrolizumab outcome—that can be accurately recapitulated using clinically accessible multiplexed IHC (mIHC) platforms. The results underscore a key determinant of spatial cellular organization, namely a distancing balance of effector and suppressive T cell activity, for predicting anti-PD-1 immunotherapy response in CTCL.

## RESULTS

### CTCL clinical cohort and multimodal experimental approach

We analyzed pre- and post-treatment biopsies from 14 patients with advanced-stage CTCL, who received pembrolizumab every 3 weeks for up to 2 years as part of the CITN-10 trial (NCT02243579) (**Fig. 1a.1, Sup. Fig. 1a-b, Sup. Table 1**)^13^. The post-treatment biopsies were collected at several timepoints, which is detailed per patient in **Sup. Fig. 1c** and **Sup. Table 1**. The spatial organization and immunogenomic heterogeneity of the CTCL TME was dissected using a formalin-fixed paraffin-embedded (FFPE) tissue microarray from 70 patient-matched pre- and post-treatment skin tumor samples (**Fig. 1a.2**). The tissue microarray spots were selected from the most infiltrated regions of the skin biopsies. CODEX multiplexed protein imaging identified 117,170 cells in the tissue microarray. These results were integrated with 64 tissue transcriptomes obtained from serial sections using laser capture microdissection and Smart-3Seq^21^ (i.e., RNA-seq) (**Fig. 1a.3**). Integrative analyses, including cellular neighborhood assessment^19^ and CIBERSORTx (CSx)^22^, were then used to profile the molecular dynamics of CTCL and identify predictive biomarkers of anti-PD-1 immunotherapy (**Fig. 1a.4**).

**Fig 1.**
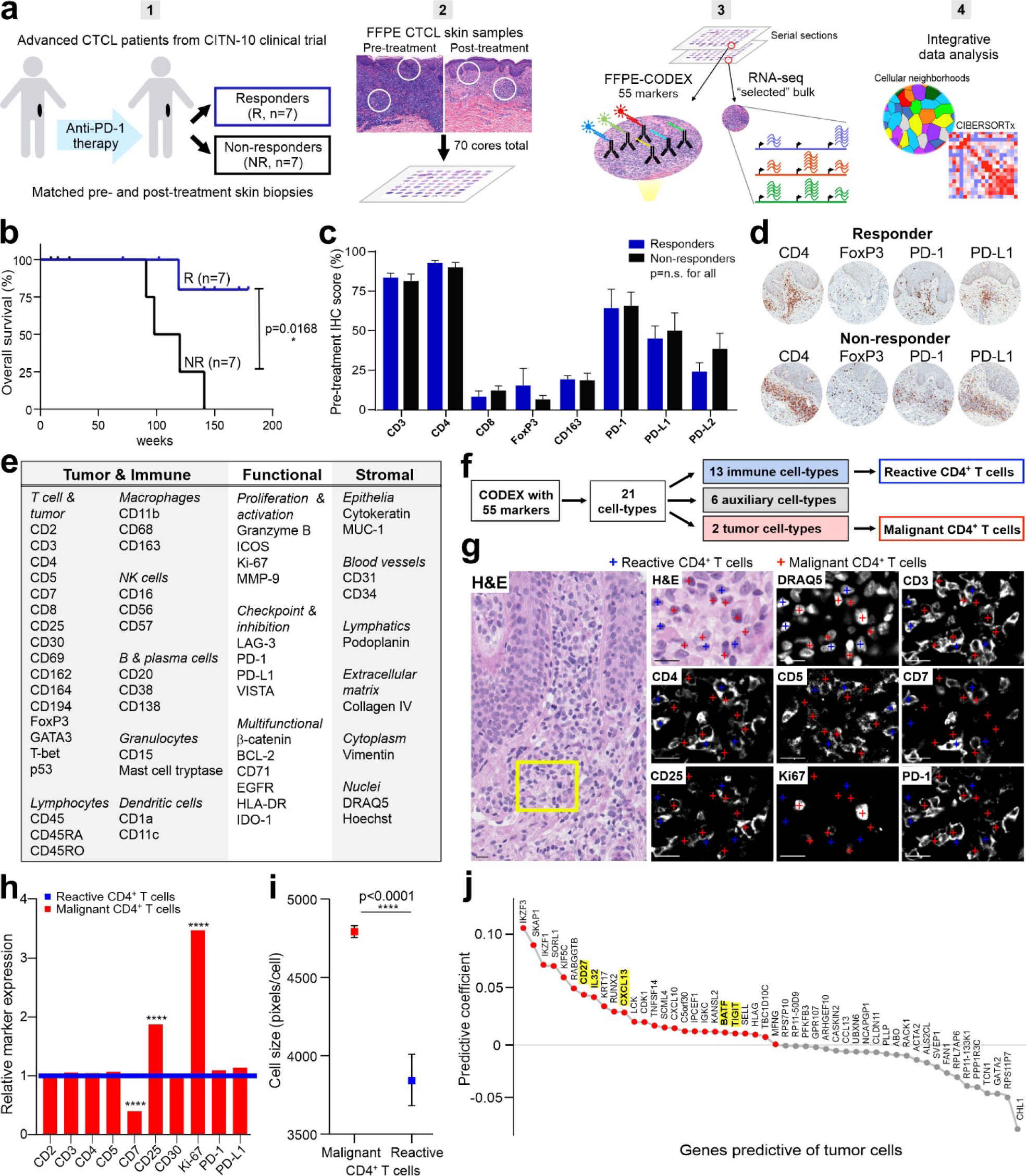
Discrimination of malignant and reactive CD4^+^ T cells in the CTCL TME. **a**, Workflow for sample preparation, CODEX, RNAseq, and computational analyses. **b**, Kaplan-Meier overall survival curve, comparing responders (R, *n*=7, blue line) and non-responders (NR, *n*=7, black line) (hazard ratio 0.0969 responder/non-responder; *p* value calculated by log-rank test). **c**, IHC protein marker expression in responders and non-responders pre-treatment (mean ± s.e.m.). *P* values were calculated by two-sided Wilcoxon’s rank-sum tests (*p* = not significant (n.s.) for all comparisons). **d**, Representative pre-treatment IHC images for select markers from a responder (top) and non-responder (bottom). **e**, CODEX antibody panel (see also **Sup. Fig. 1d**). **f**, Identification of 21 cell-types by clustering (see also **Sup. Fig. 2a-b**). **g**, Visual verification of reactive (blue crosses) versus malignant (red crosses) CD4^+^ T cells in CTCL tissue. Scale bars, 20 µm. **h**, Average expression of select markers on malignant (red bars) relative to reactive (blue line) CD4^+^ T cells. *P* values were calculated by two-sided Wilcoxon’s rank-sum tests (****, *p*<0.0001). **i**, Cell size, measured in pixels/cell, of all malignant (red square) versus reactive (blue square) CD4^+^ T cells (mean ± s.e.m.). *P* value was calculated by a two-sided Wilcoxon’s rank-sum test. **j**, Ranking genes most predictive of tumor cells per tissue microarray spot using an L1-regularized linear model. Red colored genes have positive predictive coefficients (i.e., most likely to represent tumor cells); gray colored genes have negative predictive coefficients (i.e., less likely to represent tumor cells). Known CTCL marker genes are highlighted in yellow.

Therapeutic response to pembrolizumab was assessed by consensus global response criteria^23^. No significant differences were observed at baseline between responders and non-responders for patient demographics, cancer type/stage, and treatment history (**Sup. Fig. 1b**). Clinical outcomes were significantly different between patient groups (**Sup. Fig. 1b**). Responders had a significant improvement in their overall skin response compared to non-responders, as measured by the modified Severity Weighted Assessment Tool (mSWAT)^24^ (**Sup. Fig. 1b**). Similarly, overall survival was significantly longer in responders than non-responders: Non-responders had a median survival of 109 weeks after treatment initiation, whereas all but one responder was alive at the median follow-up time of 142 weeks (**Fig. 1b**). The expression of key T cell, macrophage and PD-1 signaling markers was assessed by standard IHC for each patient’s pre-treatment tumor biopsy: PD-L2 expression was slightly increased in non-responders compared to responders but did not reach statistical significance (**Fig. 1c**). No differences were observed for the other seven markers, as shown for CD4, FoxP3, PD-1, and PD-L1 (**Fig. 1d**).

### Discrimination of malignant from reactive CD4^+^ T cells in the CTCL TME

Distinguishing malignant from reactive CD4^+^ T cells is a major challenge in CTCL, because no protein marker is 100% specific for tumor cells and both cell-types can be clonal^25^. Using a 55-marker CODEX panel (**Fig. 1e, Sup. Fig. 1d, Sup. Table 2**), unsupervised machine learning and manual curation based on marker expression, tissue localization, and morphology, we identified and validated 21 unique cell-type clusters (**Fig. 1f, Sup. Fig. 2a-b**), including reactive CD4^+^ T cells and malignant CD4^+^ T cells (i.e., tumor cells). Comparing the fluorescent stainings of reactive CD4^+^ T cells (blue crosses) and malignant CD4^+^ T cells (red crosses) showed that in tumor cells CD7 was decreased and CD25 and Ki-67 were increased (**Fig. 1g**), consistent with an advanced CTCL phenotype^26^. Quantifying these expression differences for tumor cells relative to reactive CD4^+^ T cells showed a fold-change of 0.40 for CD7, 1.87 for CD25, and 3.47 for Ki-67 (**Fig. 1h**, *p*<0.0001). Tumor cells were also larger in size than reactive CD4^+^ T cells (**Fig. 1i**), in line with previous reports^27^. One patient had large cell transformation, but the average size of tumor cells in that patient was not significantly different from the other 13 patients. Additionally, we identified genes predictive of the frequency of tumor cells per tissue microarray spot by fitting an L1-regularized linear model to bulk RNA-seq data. This confirmed that higher expression of several known CTCL marker genes, including *CD27, IL-32, CXCL13, BATF*, and *TIGIT*^28-32^, was associated with spots with higher frequencies of tumor cells (**Fig. 1j**, see yellow highlighted gene names). Notably, CTCL tumor cells can also express FoxP3^33^, as was the case for one patient in this clinical trial cohort. Our clustering approach identified this population of malignant FoxP3^+^ CD4^+^ T cells, which differed significantly from Tregs, with lower CD3, CD4, and FoxP3 marker expression and larger cell size (**Sup. Fig. 1e**). Given the heterogeneity of CTCL, the marker expression profile of tumor cells may differ between cohorts, especially those that were heavily pre-treated. However, these findings demonstrate that multiplexed imaging can discriminate malignant from reactive CD4^+^ T cells at the single-cell level, which is supported by cell size measurements and gene expression profiling.

**Fig 2.**
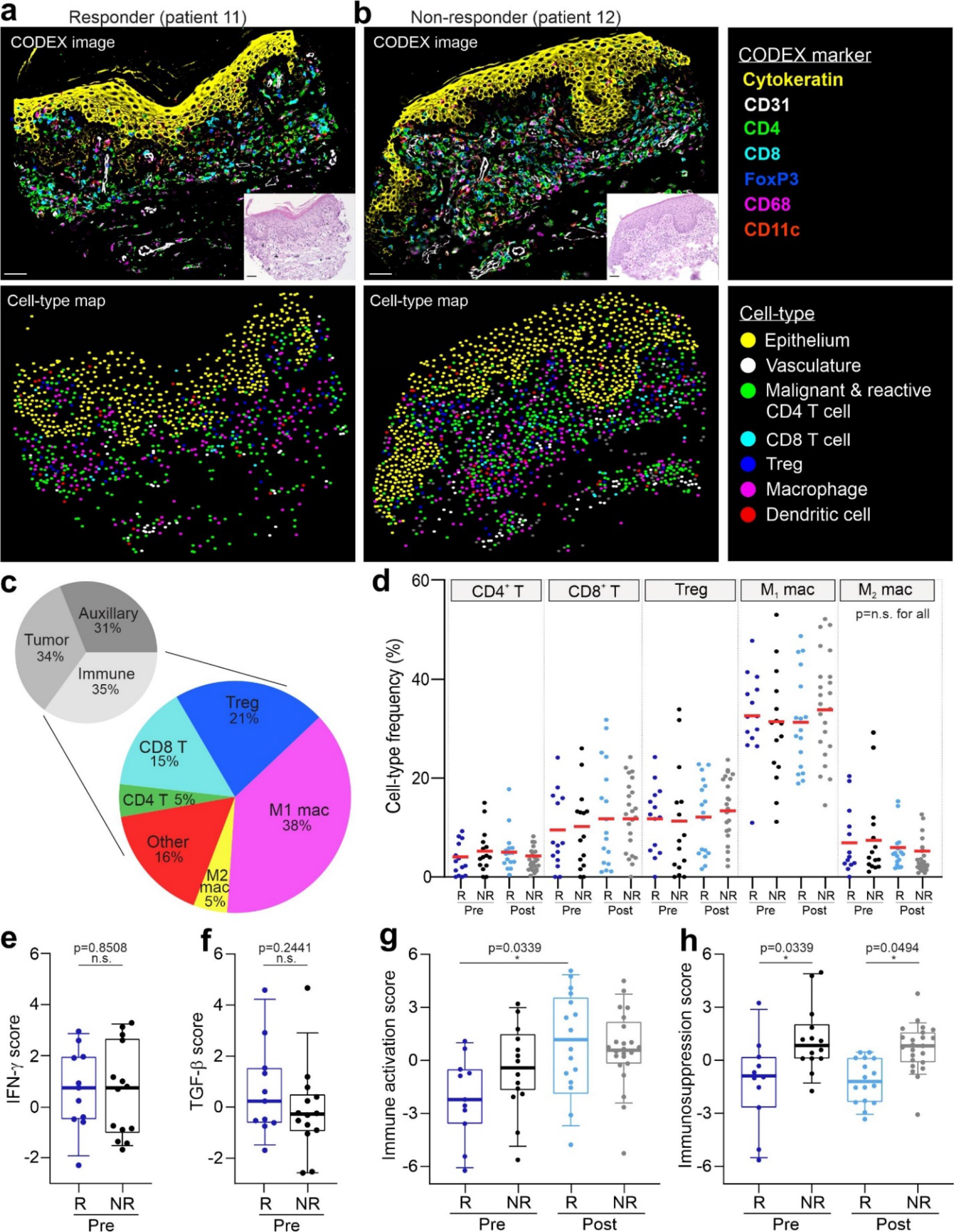
Characterization of the CTCL TME pre- and post-pembrolizumab treatment. **a-b**, Top panels: Representative CODEX seven-color overlay images from a responder (left) and non-responder (right) pre-treatment. Scale bar, 50 µm. Insets, corresponding H&E images; scale bars, 50 µm. Bottom panels: corresponding cell-type maps. **c**, Upper pie chart: overall frequencies of tumor, immune and auxiliary cell-types. Lower pie chart: overall frequencies of all immune cell-types, including CD4^+^ T cells, CD8^+^ T cells, Tregs, M1 macrophages, M2 macrophages, and other (B cells, dendritic cells, Langerhans cells, mast cells, neutrophils, and plasma cells). **d**, Cell-type frequencies of CD4^+^ T cell, CD8^+^ T cell, Treg, M1 macrophage, and M2 macrophage as a percentage of all immune cells per tissue microarray spot in responders and non-responders pre- and post-treatment (mean, red bar). *P* values were calculated with a linear mixed-effect model with Bonferroni’s corrections for multiple comparisons (*p* = not significant (n.s.) for all comparisons). **e-f**, IFN-γ (**e**) and TGF-β (**f**) gene scores between responders and non-responders pre-treatment per spot. Boxes, median ± interquartile range (IQR); whiskers, 1.5x IQR. *P* values were calculated with a linear mixed-effect model with Bonferroni’s corrections for multiple comparisons. **g-h**, Immune activation (**g**) and immunosuppression (**h**) gene scores, computed on bulk RNA-seq data, between responders and non-responders pre- and post-treatment per spot. Boxes, median ± IQR; whiskers, 1.5x IQR. *P* values were calculated with a linear mixed-effect model with Bonferroni’s corrections for multiple comparisons.

### Deep profiling of the CTCL TME in response to anti-PD-1 immunotherapy

Anti-PD-1 immunotherapies are premised on the ability to alter the delicate balance of positive and negative immune signaling to promote the recognition and elimination of tumor cells. Tumor cells, however, shift this balance in favor of their own survival and proliferation. We reasoned that delineating the complexities of tumor and immune cell interactions within the TME may reveal the cell specific organizations that determine pembrolizumab response and resistance in CTCL.

We first explored how cellular composition contributes to the efficacy of PD-1 blockade in CTCL by cataloging the frequencies of the 21 cell-types identified by CODEX across patient groups. The 21 cell-types included 13 immune cell-types, 6 auxiliary cell-types, and 2 tumor cell-types (**Sup. Fig. 2b**). Markers for T cell subsets (CD4, CD8, FoxP3), macrophages (CD68), dendritic cells (CD11c), tumor cells (CD4), vasculature (CD31), and epithelium (cytokeratin) were clearly visualized in the CODEX fluorescent images (**Fig. 2a-b, upper panels; Sup. Fig. 3a**). The corresponding hematoxylin and eosin (H&E) images (**Fig. 2a-b**, inserts; **Sup. Fig. 3b**) confirmed accurate staining of structural elements like epithelium and vasculature. Fluorescent staining of immune and tumor cells confirmed the cell-type assignments shown in the corresponding cell-type maps (**Fig. 2a-b, lower panels)**.

**Fig 3.**
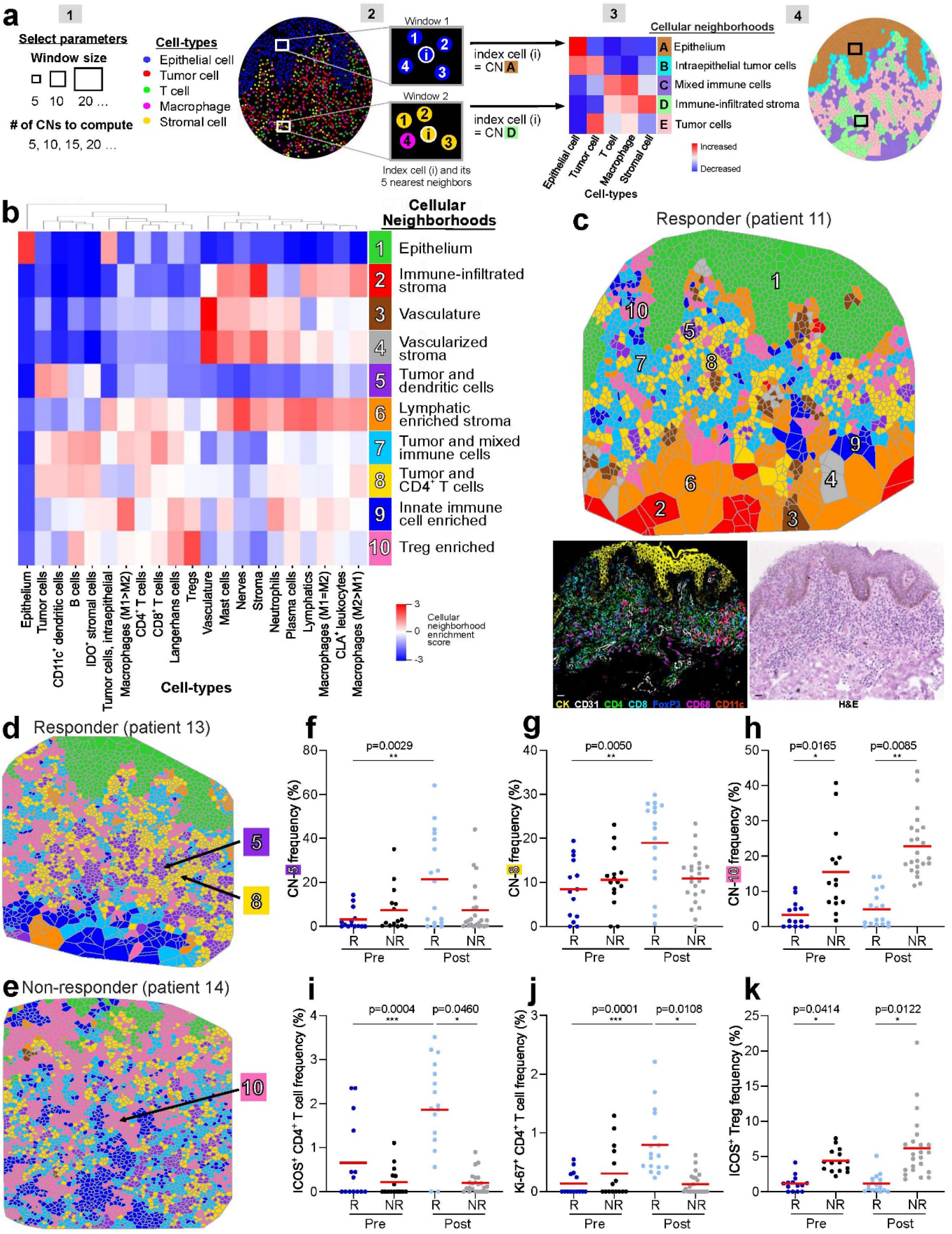
Cellular neighborhoods reveal differences in the spatial TME organization in responders and non-responders. **a**, Cellular neighborhood (CN) analysis schematic. [1] Selection of computational parameters, including the window size (five in this schematic) and the number of CNs to be computed (five in this schematic). [2] Assignment of an index cell (i, center of window) to a given CN based on the composition of cell-types within its corresponding window the clustering of windows. [3] Heatmap of cell-type distribution for each CN and assignment of CN names. [4] Visualization of CNs as a Voronoi diagram. **b**, Identification of 10 conserved CNs in the CTCL TME using a window size of 10. **c**, Representative Voronoi diagram of the 10 CNs in a responder post-treatment, with the corresponding H&E and seven color fluorescent CODEX images. Scale bar, 20 µm. **d-e**, Voronoi diagrams of CNs in a responder (**d**) and non-responder (**e**) post-treatment, highlighting CN-5 (tumor and dendritic cells), CN-8 (tumor and CD4^+^ T cells) and CN-10 (Treg enriched). **f-h**, Frequencies of CN-5 (**f**), CN-8 (**g**) and CN-10 (**h**) per tissue microarray spot in responders and non-responders pre- and post-treatment (mean, red bar). *P* values were calculated with a linear mixed-effect model with Bonferroni’s corrections for multiple comparisons. **i-k**, Frequencies of ICOS^+^ CD4^+^ T cell (**i**), Ki-67^+^ CD4^+^ T cell (**j**) and ICOS^+^ Treg (**k**) as a percentage of all immune cells per spot in responders and non-responders pre- and post-treatment (mean, red bar). *P* values were calculated with a linear mixed-effect model with Bonferroni’s corrections for multiple comparisons.

Interestingly, no differences in the content of the 21 identified cell-types were noted between responders and non-responders pre- or post-treatment (**Fig. 2c-d; Sup. Fig. 2c**). The combined frequencies of tumor, immune and auxiliary cell-types each comprised approximately one-third of all cells (**Fig. 2c, upper panel**); the same trend held across patient groups (**Sup. Fig. 2c**). Tumor cell content (averaged across all TMA spots in a patient group) was unchanged in responders or non-responders following pembrolizumab therapy (**Sup. Fig. 2c**), likely because only heavily infiltrated skin tumor biopsy regions were used for this study. Among all immune cells, the ranked cell-type frequencies were 38% for M_1_ macrophages, 21% for Tregs, 15% for CD8^+^ T cells, 5% for M2 macrophages, 5% for CD4^+^ T cells, and <5% for other immune cell-types including B cells, plasma cells, dendritic cells, Langerhans cells, mast cells, and neutrophils (**Fig. 2c, lower panel**). No differences in the mean frequencies of immune cell-types were observed when comparing responders and non-responders pre- and post-treatment; this is highlighted for T cells (CD4^+^ T, CD8^+^ T, Tregs) and macrophages (M1 and M2) (**Fig. 2d**). This finding is consistent with the initial publication on this CTCL cohort and our baseline IHC data (**Fig. 1c-d**), which showed no correlation between pembrolizumab response and the expression of T cell, macrophage or PD-1 signaling markers^13^. However, it contrasts with some solid tumor studies, which have correlated the density of tumor infiltrating lymphocytes with clinical outcome^34,35^ and immunotherapy response^36,37^.

We next focused on differences in immune signaling between responders and non-responders. Immunogenomic analyses were preformed to characterize the functional immune state of the TME, which has been shown to be a key determinant of immunotherapeutic activity^38^. Gene expression signatures that have predicted PD-1/PD-L1 blockade response (e.g., IFN-γ scores^39^, **Sup. Table 3**) and non-response (e.g., TGF-β scores^40^, **Sup. Table 3**) in solid tumors were applied to our CTCL data. No differences were observed pre-treatment in responders versus non-responders for the IFN-γ (**Fig. 2e**, median R, pre (0.77) versus NR, pre (0.76)) or TGF-β (**Fig. 2f**, median R, pre (0.28) versus NR, pre (−0.32)) gene scores; hence, neither signature was predictive of pembrolizumab response in CTCL. Since these predictive gene signatures were derived from single-disease studies, their utilization may be limited to similar solid tumor types^41^.

Rare cancers like CTCL do not have specifically defined immune gene signatures, making it a challenge to catalogue their tumor immunogenicity. Gene lists of immune activation (e.g., *CD27, EOMES*, and *ICOS*) and immunosuppression (e.g., *ENTPD1, TGFB1*, and *TIGIT*) molecules were therefore compiled (**Sup. Table 3**) and used to assess how the functional immune state of the CTCL TME was altered in response to pembrolizumab. Notably, genes like *PDCD1* and *TNFRSF18*, which can be both immune activating and suppressive depending on cellular state and microenvironmental context, were excluded from this analysis. The immune activation gene score was significantly increased in responders post-treatment compared with pre-treatment (**Fig. 2g**, median R, pre (−2.54) versus R, post (1.21)), with no significant change in non-responders post-versus pre-treatment. This indicates that only responders develop an immune activated TME phenotype following pembrolizumab therapy. In contrast, the immunosuppression gene scores were significantly increased in non-responders compared to responders both pre- and post-treatment (**Fig. 2h**, median NR, pre (0.59) versus R, pre (−0.83) and NR, post (0.64) versus R, post (−1.12)), implying that non-responders have an immunosuppressed TME phenotype relative to responders. Collectively, these results demonstrate that the functional immune state of the CTCL TME depends on factors beyond raw immune cell counts. Exploring the spatial patterns of immune infiltration is an essential step towards understanding the indicators predictive of immunotherapy outcome.

### PD-1 blockade induces spatial re-organization of the CTCL TME

It is expected that the functional immune state of the TME will be mediated by its spatial cellular organization. Cellular neighborhood (CN) analysis, applied previously in a study on colorectal cancer^19^, was performed to obtain a high level view of CTCL tissue architecture. CNs are analogous to urban neighborhoods—essentially geographically localized areas within a larger city that facilitate social interaction^42^ (**Sup. Fig. 2d**). Likewise, CNs are defined by a localized enrichment of specific cell-types within the tissue that mediate cellular interactions and vital tissue functions (**Sup. Fig. 2e**). Computationally, the CN algorithm extracts quantitative data on the composition and spatial distribution of individual cells to reveal how local cellular niches are organized within tissues (**Fig. 3a**)^19^. Specifically, computational parameters like the window size and number of CNs to be computed are manually set (**Fig. 3a.1**). Each cell in the tissue is assigned to a given CN based on the composition of cell-types within the specified window (**Fig. 3a.2**). The windows are then clustered and the correlation of CNs and cell-types are represented as a heatmap (**Fig. 3a.3**). CNs are visualized as Voronoi diagrams and analyzed to better understand cellular spatial behavior (**Fig. 3a.4**).

Using a window size of 10 nearest neighbors, we identified 10 distinct CNs that were conserved across this CTCL cohort (**Fig. 3b, Sup. Fig. 3c**). Some CNs recapitulated structural components that are clearly discernable in the corresponding H&E and fluorescent images, such as epithelium (CN-1, green region) and vasculature (CN-3, brown region) (**Fig. 3c**). The other CNs were composed of previously unappreciated sub-structures within the dermal infiltrate that were not apparent in the corresponding H&E or fluorescent images, including immune-infiltrated stroma (CN-2, red region), vascularized stroma (CN-4, gray region), tumor and dendritic cells (CN-5, purple region), lymphatic enriched stroma (CN-6, orange region), tumor and mixed immune cells (CN-7, cyan region), tumor and CD4^+^ T cells (CN-8, yellow region), innate immune cell enriched (CN-9, blue region), and Treg enriched (CN-10, pink region) (**Fig. 3c**). Representative Voronoi diagrams from a responder (**Fig. 3d**, see CN-5 and CN-8, purple and yellow regions, respectively) and non-responder (**Fig. 3e**, see CN-10, pink region) post-treatment show significant differences in the frequency of CNs between patient groups. CNs enriched in tumor and dendritic cells (CN-5, purple region) (**Fig. 3f**, mean frequency in R, post (21.4%) versus R, pre (3.2%)) and tumor and CD4^+^ T cells (CN-8, yellow region) (**Fig. 3g**, mean frequency in R, post (19.0%) versus R, pre (8.5%)) were present at significantly higher levels in responders post-treatment than in non-responders or either group pre-treatment. In contrast, the Treg enriched CN (CN-10, pink region) was present at a significantly higher frequency in non-responders than responders pre- and post-treatment (**Fig. 3h**, mean frequency in NR, pre (15.5%) versus R, pre (3.3%) and NR, post (22.7%) versus R, post (4.9%)). Notably, these differences in the spatial organization of certain cell-types were observed even though there were no differences in the abundance of CD4^+^ T cells, dendritic cells, Tregs, or tumor cells between patient groups, as previously described (**Fig. 2c-d**).

The increased co-localization of dendritic cells, CD4^+^ T cells and tumor cells in responders post-treatment, relative to other patient groups, suggests that the more immune activated TME observed in responders following pembrolizumab therapy (i.e., increased immune activation gene score in **Fig. 2g**) may in part be mediated by CD4^+^ T cell activation by antigen-presenting cells. Indeed, after PD-1 blockade, responders had increased frequencies of activated ICOS^+^ CD4^+^ T cells (**Fig. 3i**, mean frequency in R, post (1.9%) versus R, pre (0.7%) and NR, post (0.2%)) and proliferating Ki-67^+^ CD4^+^ T cells (**Fig. 3j**, mean frequency in R, post (0.8%) versus R, pre (0.1%) and NR, post (0.1%)) compared to responders pre-treatment and non-responders post-treatment, suggesting that pembrolizumab therapy activates CD4^+^ T cells through upregulation of immune activating molecules.

The increased frequency of the Treg enriched CN in non-responders pre- and post-treatment is consistent with our finding that non-responders have a persistently immunosuppressed TME (i.e., increased immunosuppression gene score in **Fig. 2h**), which is further supported by the higher frequency of a potently suppressive subset of Tregs expressing ICOS in non-responders (**Fig. 3k**, mean frequency in NR, pre (4.4%) versus R, pre (1.2%) and NR, post (6.2%) versus R, post (1.2%))^43,44^. Interestingly, despite treatment with PD-1 blockade, no differences were observed in PD-1^+^ subsets of CD4^+^, CD8^+^, Tregs, or tumor cells between groups (**Sup. Fig. 2-i**). Collectively, these data link the spatial re-organization of CD4^+^ T cells, tumor cells, and Tregs with immune activity of the TME, highlighting patterns of immune control and pembrolizumab responsiveness.

### Spatial signature of PD-1^+^ CD4^+^ T cells, tumor cells, and Tregs predicts pembrolizumab response in CTCL

It is reasonable to suspect that specific cellular interactions drive pembrolizumab responsiveness in CTCL. While complex spatial analyses have been used to prognosticate cancer outcomes in select clinical cohorts^19^, we hypothesized that a simplified approach based on the distances between specific immune and tumor cells could be employed. Guided by our CN findings, which highlighted the spatial arrangement of CD4^+^ T cells, tumor cells and Tregs, we formalized a computational scoring approach to standardizing a spatial signature. This score, termed *SpatialScore*, calculates the physical distance ratio of each CD4^+^ T cell and its nearest tumor cell (“right” distance) relative to its nearest Treg (“left” distance) (**Fig. 4a**). A lower *SpatialScore* (i.e., CD4^+^ T cells are closer to tumor cells than Tregs) suggests increased T cell effector activity (**Fig. 4a.1**), whereas a higher *SpatialScore* (i.e., CD4^+^ T cells are closer to Tregs than tumor cells) suggests increased T cell suppression (**Fig. 4a.2**). As such, the *SpatialScore* can be viewed as a proxy of the balance between T cell effector activity and suppression in the TME.

**Fig 4.**
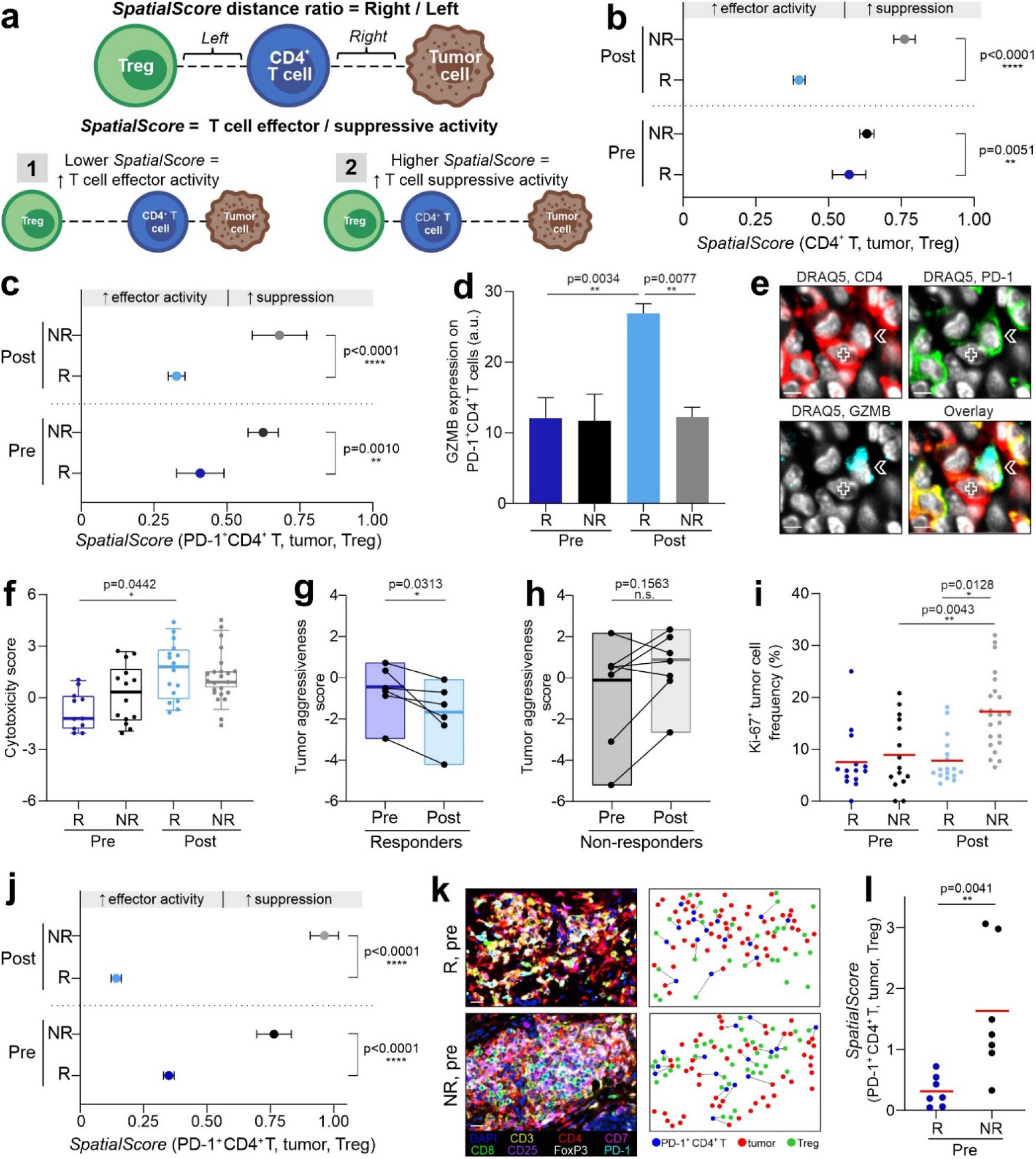
Spatial relationship between CD4^+^ T cells, Tregs and tumor cells predicts pembrolizumab response in CTCL. **a**, *SpatialScore* schematic. The *SpatialScore* is calculated by taking the ratio of the physical distance between each CD4^+^ T cell and its nearest tumor cell (distance “right”) relative to its nearest Treg (distance “left”). [1] A lower *SpatialScore* (i.e., CD4^+^ T cells are closer to tumor cells than Tregs) suggests increased T cell effector activity. [2] A higher spatial score (i.e., CD4^+^ T cells are closer to Tregs than tumor cells) suggests increased T cell suppression. **b-c**, *SpatialScore* on a per cell basis for CD4^+^ T cells (**b**) and PD-1^+^ CD4^+^ T cells (**c**) for each patient group (mean ± s.e.m.). *P* values were calculated by two-sided Wilcoxon’s rank-sum tests, with no adjustments for multiple comparisons. **d**, GZMB protein expression on PD-1^+^ CD4^+^ T cells by CODEX in responders and non-responders pre- and post-treatment per tissue microarray spot (mean fluorescence intensity (arbitrary units, a.u.) ± s.e.m.) *P* values were calculated with a linear mixed-effect model with Bonferroni’s corrections for multiple comparisons. **e**, CODEX images of DRAQ5 (white), CD4 (red), PD-1 (green), GZMB (cyan), and overlay showing contact between a tumor cell (marked by cross) and GZMB-expressing PD-1^+^ CD4^+^ T cell (marked by arrow) in responder patient 13 post-treatment. Scale bars, 10 µm. **f**, Cytotoxicity gene scores, computed on bulk RNA-seq data, between responders and non-responders pre- and post-treatment per spot. Boxes, median ± IQR; whiskers, 1.5x IQR. *P* values were calculated with a linear mixed-effect model with Bonferroni’s corrections for multiple comparisons. **g-h**, Pre-to post-treatment changes in tumor aggressiveness gene scores, computed on bulk RNA-seq data, on a per patient basis in responders (**g**) and non-responders (**h**). Boxes, median ± IQR; whiskers, 1.5x IQR. *P* values were calculated by two-sided Wilcoxon’s signed-rank tests. **i**, Ki-67^+^ tumor cell frequencies per spot in responders and non-responders pre- and post-treatment (mean, red bar). *P* values were calculated with a linear mixed-effect model with Bonferroni’s corrections for multiple comparisons. **j**, *SpatialScore* on a per cell basis calculated from Vectra mIHC data using PD-1^+^CD4^+^ T cells for each patient group (mean ± s.e.m.). *P* values were calculated by two-sided Wilcoxon’s rank-sum tests, with no adjustments for multiple comparisons. **k**, Vectra mIHC images (left panels; representative regions of a tissue microarray spot) in responder patient 13 (R) and non-responder patient 14 (NR) pre-treatment. Scale bars, 20 µm. The corresponding spatial plots (right panels) show that PD-1^+^ CD4^+^ T cells (blue dots) are closer to tumor cells (red dots) than Tregs (green dots) in the responder and vice versa in the non-responder. **l**, Pre-treatment *SpatialScore* calculated from Vectra mIHC data for responders and non-responders on a per patient basis (mean, red bar). *P* values was calculated by a two-sided Wilcoxon’s rank-sum test.

The *SpatialScore* is calculated on a per cell basis and the mean value is reported for each patient group. When calculated with all CD4^+^ T cells, the pre-treatment *SpatialScore* was significantly lower in responders than non-responders, with enhancement post-treatment for both groups (**Fig. 4b**, compare mean *SpatialScore* for R, pre (0.57) versus NR, pre (0.63)). The same trend was observed on a per patient basis (**Sup. Fig. 4a**). Since the current study trialed PD-1 blockade in CTCL, we next asked how the *SpatialScore* was influenced by the PD-1^+^ CD4^+^ T cell subset. As observed when calculated with all CD4^+^ T cells, when computed with PD-1^+^ CD4^+^ T cells, tumor cells, and Tregs, the *SpatialScore* was lower in responders than non-responders (**Fig. 4c**, compare mean *SpatialScore* for R, pre (0.41) versus NR, pre (0.62)). The same trend was seen on a per patient basis (**Sup. Fig. 4b**). Notably, the *SpatialScore* was lower in responders pre-treatment when calculated with PD-1^+^ CD4^+^ T cells versus all CD4^+^ T cells, implying increased effector activity in this T cell subset. These results suggest that PD-1^+^ CD4^+^ T cells in responders are primed for increased anti-tumor activity, which is enhanced in the immune activated TME that results following pembrolizumab therapy.

Since the *SpatialScore* approach appears to predict the outcome of PD-1 blockade in CTCL, it stands to reason that there is a deep phenotype of cell-type specific architecture that is driving the *SpatialScore*. Pre-treatment differences were driven by the closer proximity of PD-1^+^ CD4^+^ T cells and Tregs in non-responders (**Sup. Fig. 4c**, see red arrow), consistent with the increased immunosuppression gene scores in non-responders relative to responders (**Fig. 2h**). In contrast, post-treatment differences were driven by the closer proximity of PD-1^+^ CD4^+^ T cells and tumor cells in responders (**Sup. Fig. 4d**, see red arrow), consistent with the increased immune activation gene score in responders relative to non-responders (**Fig. 2i**). Importantly, no correlation was identified between the abundance of PD-1^+^ CD4^+^ T cells, tumor cells, or Tregs and the *SpatialScore* on a per tissue microarray spot basis (**Sup. Fig. 4g-i**), implying that the *SpatialScore* is not merely driven by cell-type frequency. Additionally, the mean *SpatialScore* was significantly different from that of a random sample in responders pre- and post-treatment (**Sup. Fig. 4j and l**), but not in non-responders pre- and post-treatment (**Sup. Fig. 4k and m**).

This suggests that an active process coordinates the spatial interactions of PD-1^+^ CD4^+^ T cells, tumor cells and Tregs in responders, whereas no such mechanism exists in non-responders. Finally, when the *SpatialScore* was calculated for CD8^+^ T cells (**Sup. Fig. 4e**) or PD-1^+^ CD8^+^ T cells (**Sup. Fig. 4f**), it was not predictive of pembrolizumab response. The increased proximity of CD4^+^ T cells and tumor cells observed in pembrolizumab responders—and lack thereof for CD8^+^ T cells—suggests that CD4^+^ T cells may have a more important effector function in CTCL than previously appreciated.

### PD-1^+^ CD4^+^ T cells upregulate cytotoxic effector molecules in pembrolizumab responders

We reasoned that the spatial proximity of PD-1^+^ CD4^+^ T cells and tumor cells reflected the enhanced CD4^+^ T cell tumor reactivity seen in responders following pembrolizumab therapy. CODEX and RNA-seq datasets were therefore used to assess the cytotoxic potential of effector PD-1^+^ CD4^+^ T cells. In CTCL, cytotoxic CD4^+^ T cells act through a granzyme-perforin-dependent pathway to achieve tumor cell killing^45,46^, similar to cytotoxic CD8^+^ T cells^46-48^. The expression of granzyme B (GZMB) on PD-1^+^ CD4^+^ T cells was increased in responders post-versus pre-treatment (**Fig. 4d**, mean GZMB expression, R, post (26.9) versus NR, post (12.2) and R, pre (12.1)). This increased GZMB expression is confirmed visually in a responder post-treatment with DRAQ5 (nuclear stain), CD4, PD-1, and GZMB fluorescent staining (**Fig. 4e**, see white arrow) and is consistent with the increased cytotoxicity gene score^49,50^ seen in responders post-treatment compared to pre-treatment (**Fig. 4f**, median R, post (1.21) versus R, pre (−1.77), **Sup. Table 3**). No differences were observed between non-responders pre- and post-treatment with regards to GZMB expression on PD-1^+^ CD4^+^ T cells (**Fig. 4d**, mean expression, NR, pre (11.7) versus NR, post (12.2)) or the cytotoxicity gene score (**Fig. 4e**, median NR, pre (0.21) versus NR, post (0.43)).

Increased cytotoxicity should coincide with decreased tumor cell aggressiveness and, using genes associated with poor response to therapy^51^ and progressive^52^ CTCL (**Sup. Table 3**), it was observed that tumor aggressiveness decreased in 100% of responders following pembrolizumab therapy (**Fig. 4g**) but increased in 71% of non-responders (**Fig. 4h**). This increased tumor aggressiveness in non-responders was further supported by an increased frequency of proliferating (Ki-67^+^) tumor cells in non-responders post-versus pre-treatment and compared to responders post-treatment (**Fig. 4i**, mean frequency, NR, post (17.0%) versus NR, pre (8.9%) and R, post (7.8%))^26,53^. Collectively, these data show the cytotoxic potential of effector PD-1^+^ CD4^+^ T cells and their association with a favorable response to pembrolizumab therapy in CTCL.

### Validation of the *SpatialScore* biomarker with a clinically accessible multiplexed IHC platform

While mIHC approaches, such as CODEX, provide the raw data for deep cellular profiling, translating these findings to a clinical arena requires simplifying the predictive signature to a diagnostic platform that can be readily implemented in clinical practice. Vectra, in conjugation with the Phenoptics workflow, is a commercially available, widely adopted clinical mIHC imaging platform, which has been used to identify biomarkers for renal cell carcinoma^54^ and B cell lymphoma^55^. We devised a simplified staining panel (DAPI, CD3, CD4, CD7, CD8, CD25, FoxP3, and PD-1) that captured the PD-1^+^ CD4^+^ T cells, tumor cells, and Tregs used to calculate the *SpatialScore*. Notably, the tumor cell phenotype identified by CODEX (i.e., decreased expression of CD7 and increased expression of CD25 and Ki-67; **Fig. 1h**), was critical for establishing this streamlined staining panel and could be readily transferred to the Vectra platform.

Serial sections from the same CTCL TMA used for CODEX and RNA-seq were stained with this simplified panel and imaged with Vectra (**Sup. Fig. 5a**). Across all TMA spots, 126,653 cells were identified, including 2,957 PD-1^+^ CD4^+^ T cells, 6,161 Tregs, and 19,847 tumor cells. The *SpatialScore* was then computed on a per cell basis and the mean was reported for each patient group. Consistent with the CODEX results (**Fig. 4c**), the Vectra-calculated *SpatialScore* was significantly lower in responders than non-responders pre-treatment (**Fig. 4j**, mean *SpatialScore* R, pre (0.35) versus NR, pre (0.76)), with enhancement post-treatment compared to pre-treatment. The Vectra mIHC images (**Fig. 4k**, left) and corresponding spatial maps (**Fig. 4k**, right) provide visual validation at the single-cell level that PD-1^+^ CD4^+^ T cells were closer to tumor cells in responders but closer to Tregs in non-responders. Furthermore, on a per patient basis, the pre-treatment *SpatialScore* was nearly 5-times lower in responders than non-responders (**Fig. 4l**, mean *SpatialScore* R, pre (0.31) versus NR, pre (1.52)), with excellent biomarker performance measures at a *SpatialScore* cutoff point of 0.7908 (**Sup. Fig. 5b-e**). Notably, when calculated per patient, the *SpatialScore* extended over a wider range than when calculated for all cells per group; this is due to sample size differences (i.e., n=7 per patient versus n=1000s of cells per group). It is noteworthy that the skin-derived *SpatialScore* is so robust and predictive of pembrolizumab response, since clinical outcome in advanced CTCL is often driven by the burden of disease in the non-skin compartments (i.e., blood or viscera). Collectively, these results show that findings identified by highly multiplexed imaging platforms like CODEX can be translated to more simplified platforms like Vectra, further demonstrating the clinical utility of the *SpatialScore* as a biomarker predictive of PD-1 blockade outcome in CTCL.

**Fig 5.**
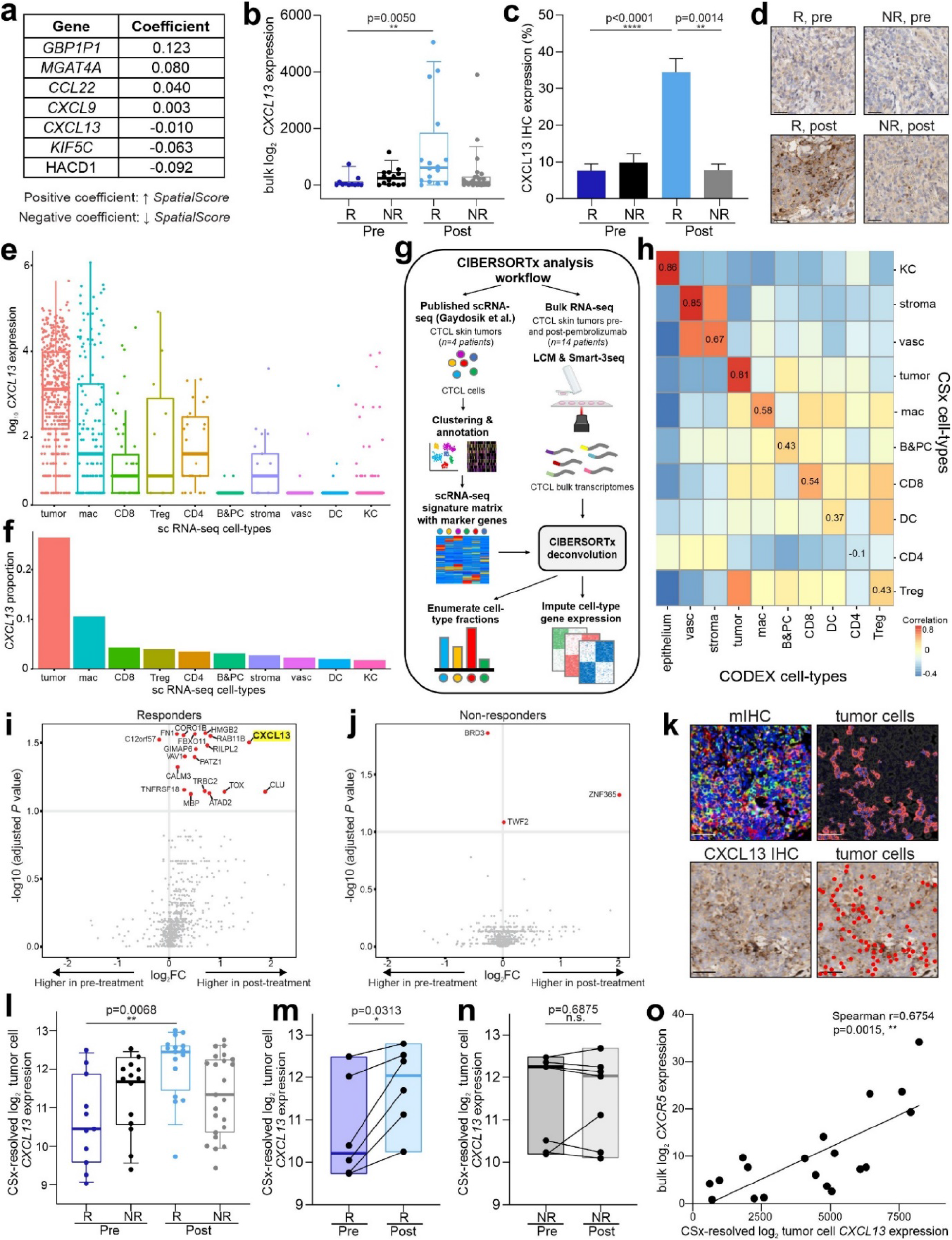
CXCL13 is a key driver of pembrolizumab response in CTCL. **a**, Seven genes predictive of the *SpatialScore* identified from bulk RNA-seq data. Genes with a positive coefficient are associated with a higher *SpatialScore* (i.e., a suppressive phenotype), whereas those with a negative coefficient are associated with a lower *SpatialScore* (i.e., an effector phenotype). **b**, Normalized bulk *CXCL13* gene expression between responders and non-responders pre- and post-treatment per tissue microarray spot. Boxes, median ± IQR; whiskers, 1.5x IQR. *P* values were calculated with a linear mixed-effect model with Bonferroni’s corrections for multiple comparisons. **c**, CXCL13 protein expression by IHC in responders and non-responders pre- and post-treatment per spot (mean ± s.e.m.). *P* values were calculated with a linear mixed-effect model with Bonferroni’s corrections for multiple comparisons. **d**, Representative CXCL13 IHC images from responder patient 9 pre-treatment (top left) and post-treatment (bottom left) as well as non-responder patient 14 pre-treatment (top right) and post-treatment (bottom right). Scale bars, 20 µm. **e-f**, Single-cell transcriptomes from a publicly available scRNA-seq dataset of CTCL skin tumors (Gaydosik et al.)^59^ were analyzed for *CXCL13* expression. Abbreviations: mac (macrophages), B&PC (B and plasma cells), vasc (vasculature), DC (dendritic cells), and KC (keratinocytes). **e**, Normalized expression of *CXCL13* in single cells (each dot represents a cell positive for *CXCL13*). Cells with *CXCL13* log1p normalized read counts less than 0.5 were excluded (see **Methods**). Boxes, median ± IQR; whiskers, 1.5x IQR. **f**, Proportion of CXCL13-expressing cells per cell-type. **g**, CIBERSORTx analysis workflow schematic. Single cell transcriptomes from Gaydosik et al.^59^ were used to generate a CSx deconvolution signature matrix, consisting of cell-type-specific marker genes (left portion of schematic). This matrix was then applied to CTCL bulk transcriptomes obtained with laser-capture microdissection (LCM) and Smart-3Seq (right portion of schematic) to enumerate cell-type fractions and resolve gene expression profiles (see **Methods**). **h**, Heatmap of the correlation between CSx-resolved cell-type frequencies and CODEX-identified cell-type frequencies. Spearman correlation coefficients between corresponding cell-types are shown along the diagonal. For this analysis, some of the original 21 CODEX cell-types were manually merged to match CSx cell-types (e.g., B cells and plasma cells). **i-j**, Volcano plots of differential gene expression of CSx-resolved tumor cell genes in responders (**j**) and non-responders (**k**) pre- and post-treatment. *P* values were calculated with a linear mixed-effect model with Benjamini-Hochberg correction. Significantly different genes (*p* < 0.1) are colored red; *CXCL13* is highlighted yellow. **k**, Vectra mIHC image of DAPI (blue), CD3 (yellow), CD4 (red), CD7 (magenta), CD8 (green), CD25 (purple), FoxP3 (white), and PD-1 (cyan) (top left), corresponding tumor cell depiction (top right), corresponding CXCL13 IHC image (bottom left), and overlaid image showing that CXCL13 IHC staining most commonly localized to tumor cells (red circles) (bottom right) in responder patient 9 post-treatment. Scale bars, 20 µm. **l**, Normalized CSx-resolved *CXCL13* expression in tumor cells between responders and non-responders pre- and post-treatment per spot. Boxes, median ± IQR; whiskers, 1.5x IQR. *P* values were calculated with a linear mixed-effect model with Bonferroni’s corrections for multiple comparisons. **m-n**, Pre-to post-treatment changes in normalized *CXCL13* gene expression from CSx-resolved tumor genes on a per patient basis in responders (**m**) and non-responders (**n**). Boxes, median ± IQR; whiskers, 1.5x IQR. *P* values were calculated by two-sided Wilcoxon’s signed-rank tests. **o**, Correlation of CSx-resolved tumor cell *CXCL13* expression and bulk *CXCR5* expression per spot. Data was evaluated with the Spearman test and a two-tailed t-distribution with n-2 degrees of freedom.

### Tumor cell-specific CXCL13 expression coincides with a favorable response to PD-1 blockade in CTCL

Potential recruitment mechanisms driving the *SpatialScore* were investigated by identifying genes predictive of the spatial interactions between PD-1^+^ CD4^+^ T cells, tumor cells, and Tregs. An L1-regularized linear model was fit to bulk RNA-seq data on a per spot basis, which revealed seven genes predictive of the *SpatialScore* including three chemokines: *CXCL9, CCL22* and *CXCL13* (**Fig. 5a**). *CXCL9* and *CCL22* are known to mediate Treg recruitment^56,57^; they had positive coefficients and were predictive of the higher *SpatialScore* seen in non-responders. *CXCL13* is a chemoattractant expressed on benign lymphocytes and CTCL tumor cells^28,58^; it had a negative coefficient and was predictive of the lower *SpatialScore* seen in responders. Bulk *CXCL13* gene expression was significantly increased in responders post-versus pre-treatment and compared to non-responders post-treatment (**Fig. 5b**, median normalized *CXCL13* expression, R, post (615.5) versus NR, post (94.7) and R, pre (16.0)). This upregulation of *CXCL13* in responders post-treatment relative to other patient groups was confirmed by CXCL13 IHC shown quantitatively (**Fig. 5c**, mean CXCL13 IHC expression, R, post (34.5) versus NR, post (7.8) and R, pre (7.6)) and visually (**Fig. 5d**, see R, post for highest expression of CXCL13).

We next analyzed a publicly available scRNA-seq dataset of four CTCL skin tumors (Gaydosik et al.)^59^ to identify the main cell-type overexpressing *CXCL13*. Using this dataset, we annotated 10 clusters including tumor cells, CD4^+^ T cells (CD4), Tregs, CD8^+^ T cells (CD8), B & plasma cells (B&PC), macrophages (mac), dendritic cells (DC), stroma, vasculature (vasc), and keratinocytes (KC) (**Sup. Fig. 6a-b**). Examining all cells with a *CXCL13* log1p normalized read count > 0.5 showed that tumor cells had the highest mean expression compared to other cell-types (**Fig. 5e**, median *CXCL13* expression, tumor (3.3) versus less than 2.0 for the other 9 cell-types). Moreover, the frequency of *CXCL13* expressing cells was highest among tumor cells compared to other cell-types (**Fig. 5f**, mean frequency of *CXCL13* positive cells, tumor (0.27) versus less than 0.1 for the other 9 cell-types).

**Fig 6.**
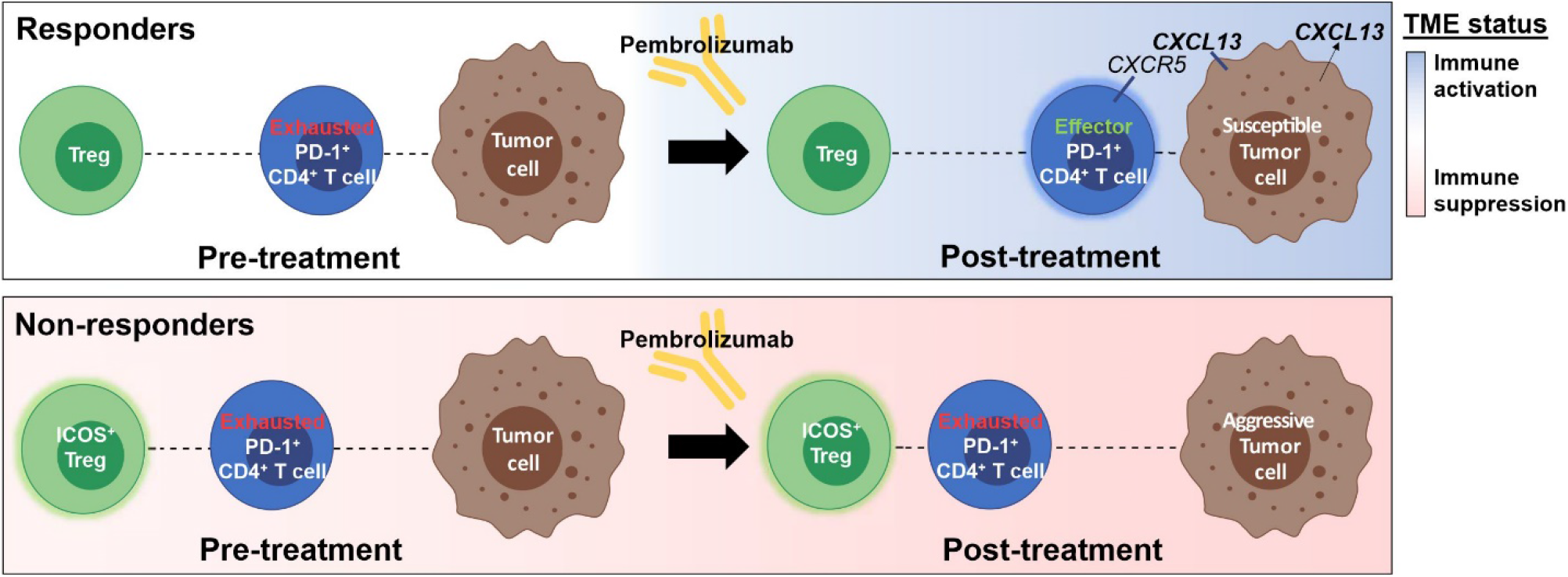
Proposed mechanisms of pembrolizumab response in CTCL. Proposed mechanisms of pembrolizumab response in therapy responders (top panel) and non-responders (bottom panel) pre- and post-treatment. The functional immune status of the TME is represented by blue shading when activated and pink shading when suppressed. In non-responders, the TME is continually immunosuppressed and persistently exhausted PD-1^+^CD4^+^ T cells are in closer proximity to potently suppressive ICOS^+^ Tregs. Due to pembrolizumab resistance, non-responder tumor cells become more aggressive following therapy. In contrast, responders have a neutral functional immune state pre-treatment, which becomes activated following pembrolizumab therapy, enabling the transition from exhausted to effector PD-1^+^ CD4^+^ T cells. Additionally, responder tumor cells are susceptible to PD-1 blockade and overexpress CXCL13. This attracts effector PD-1^+^ CD4^+^ T cells toward tumor cells, providing a mechanism for the sustained clinical response seen in responders.

CIBERSORTx (CSx) is a computational framework that uses gene expression signatures to enable cell type-specific gene expression to be inferred from bulk RNA-seq data without physical cell isolation^22^. We used CSx to computationally resolve tumor, stromal, and immune cell subsets in bulk RNA-seq data (**Fig. 5g** and **Sup. Table 4**). Using the 10 annotated clusters shown in **Fig. e-f**, a signature matrix, consisting of cell-type specific marker genes, was generated (**Sup. Fig 6c** and see **Methods**, marker genes are shown as rows). This signature matrix was used to enumerate CSx-resolved cell-type fractions and resolve gene expression profiles from CTCL bulk transcriptomes. Strong correlations were observed between most of the CSx-resolved cell-type frequencies and CODEX-identified cell-type frequencies (**Fig. 5h**, see correlation coefficients along the diagonal of the heatmap).

The CSx-resolved gene expression profiles of tumor cells were then thoroughly characterized and compared across patient groups. Interestingly, *RAB11B*, a RAS superfamily member of small GTP-binding proteins, was the only tumor cell gene that was significantly differentially expressed between responders and non-responders pre-treatment (**Sup. Fig. 6d**). This suggests an absence of tumor cell-intrinsic differences between responders and non-responders at baseline. In contrast, numerous genes, including *CXCL13*, were upregulated in tumor cells of responders post-treatment compared to pre-treatment (**Fig. 5i**, see *CXCL13* in bold). This finding is consistent with an increased susceptibility of responder tumor cells to PD-1 blockade therapy, which is supported by the decreased tumor aggressiveness score observed in responders post-treatment relative to pre-treatment (**Fig. 4g**). Interestingly, only three genes (*BRD3, TWF2* and *ZNF365*) were differentially expressed in non-responder tumor cells post-treatment compared to pre-treatment (**Fig. 5j**). This suggests that non-responder tumor cells are resistant to PD-1 blockade, which is in line with the increased tumor aggressiveness score observed in non-responders post-treatment compared to pre-treatment (**Fig. 4h**). Finally, co-staining the tissue section used for the Vectra mIHC experiment with a standard IHC anti-CXCL13 antibody, followed by co-localization of tumor cells with CXCL13-positive cells, provided visual confirmation that tumor cells are the primary expressors of CXCL13 (**Fig. 5k, Sup. Fig. 6e**).

Next, we assessed *CXCL13* expression in tumor cells and its role in recruiting reactive lymphocytes. CSx-resolved tumor cell-specific *CXCL13* expression was significantly increased in responders post-treatment compared with non-responders post-treatment and responders pre-treatment on a per tissue microarray spot basis (**Fig. 5l**, median normalized tumor cell-specific *CXCL13* expression, R, post (12.64) versus NR, post (11.14) and R, pre (10.54)). On a per patient basis, following PD-1 blockade, tumor cell-resolved *CXCL13* expression increased in 100% of responders (**Fig. 5m**) versus 29% of non-responders (**Fig. 5n**). CXCL13 exclusively binds to the chemokine receptor CXCR5, which is expressed on B cells, CD4^+^ T cells, CD8^+^ T cells, and skin-derived migratory dendritic cells^60^. Although not significant, *CXCR5* expression was increased in bulk mRNA of responders post-treatment relative to pre-treatment (**Sup. Fig. 6f**). The data lacked significant power to unmix *CXCR5* on CD4^+^ T cells by CSx; however, tumor cell-specific *CXCL13* expression was positively correlated (Spearman correlation coefficient = 0.68) with bulk mRNA *CXCR5* expression (**Fig. 5o**), suggesting a chemoattractant recruitment mechanism.

Collectively, these results indicate that PD-1 blockade distinctly alters the CTCL TME of therapeutic responders and non-responders. In responders, pembrolizumab therapy promotes immune activation and upregulates CXCL13 in tumor cells (**Fig. 6**, top panel). CXCL13 overexpression then attracts effector/cytotoxic PD-1^+^ CD4^+^ T cells to tumor cells, promoting CD4^+^ T cell-mediated tumor cell inhibition and killing. As such, the overexpression of CXCL13 in tumor cells provides a mechanism for the sustained clinical activity seen in responders. In contrast, non-responders have a persistently immunosuppressed TME, which mediates the increased interaction between inhibitory Tregs and PD-1^+^ CD4^+^ T cells and maintains this CD4^+^ T cell subset in an exhausted state (**Fig. 6**, bottom panel). In the setting of pembrolizumab resistance, non-responder tumor cells remain active and proliferative following treatment. Thus, underlying differences in the functional immune state of the TME—coupled with alterations in CD4^+^ T cell effector activity versus suppression, and tumor cell-specific CXCL13 expression— are associated with distinct spatial cellular patterns that predict the efficacy of PD-1 blockade in CTCL.

## DISCUSSION

For anti-PD-1 immunotherapies to provide maximal benefit to cancer patients, the drivers and resistors of clinical response must be identified. Traditional biomarker studies with IHC, gene expression profiling, and tumor mutational burden assays do not fully account for spatial cellular context and have imperfect correlation with immunotherapy outcomes^16^. This was true for our CTCL cohort: No pre-treatment differences were observed in immune composition, expression of PD-1/PD-L1 proteins, or IFN-γ/TGF-β gene signatures between responders and non-responders. We therefore explored alternative approaches, including spatially resolved multiplexed tissue imaging, which has been shown to significantly improve the accuracy of predicting response to PD-1 blockade in several tumor types^61^.

Multiparameter histologic analysis revealed global prognostic spatial patterns that were predictive of clinical response, including a localized enrichment of tumor and CD4^+^ T cells (CN-8) in responders and of Tregs (CN-10) in non-responders. Profiling the spatial relationships between effector PD-1^+^ CD4^+^ T cells, tumor cells, and immunosuppressive Tregs allowed us to derive the *SpatialScore*—a clinically useful biomarker that correlated strongly with pembrolizumab response in CTCL. Previously identified spatial biomarkers rely on pairwise distances (e.g., PD-1^+^ T cells and PD-L1^+^ tumor cells)^17,62,63^, whereas the *SpatialScore* accounts for the interactions between three functionally distinct cell-types. As such, the *SpatialScore* represents a novel predictive approach and provides insight into three key therapeutic determinants underlying PD-1 blockade in CTCL: 1) functional immune state of the TME, 2) T cell function, and 3) chemoattraction.

First, the *SpatialScore* captures differences in the functional immune phenotypes between responders and non-responders. Following pembrolizumab therapy, the TME of responders becomes activated, as evidenced by the increased immune activation gene score, increased frequency of ICOS^+^ and Ki-67^+^ CD4^+^ T cells, and a local enrichment of tumor cells with dendritic cells (CN-5) and CD4^+^ T cells (CN-8). These findings suggest that in responders PD-1 blockade specifically activates CD4^+^ T cells through the expression of immunostimulatory molecules and co-stimulation by antigen-presenting cells, resulting in CD4^+^ T cell proliferation^64,65^. In contrast, non-responders have a persistently immunosuppressed TME. This is supported by the increased frequency of a Treg enriched neighborhood (CN-10) and the expansion of a highly suppressive ICOS^+^ Treg subset, which has been associated with poor clinical outcomes in melanoma^43^ and liver cancer^44^. These results suggest that the increased suppressive function and spatial organization of Tregs in non-responders account for the strong immunosuppression and lack of pembrolizumab response. We speculate that the absence of this pre-treatment immunosuppression in responders enables priming and activation of CD4^+^ T cells following immunotherapy.

Second, the *SpatialScore* reflects differences in T cell function between responders and non-responders: After PD-1 blockade, T cell effector activity is restored in responders, whereas non-responders have a continually exhausted T cell phenotype. Consistent with studies of Hodgkin lymphoma^63^, bladder cancer^50^, and glioblastoma^66^, our data suggest that CD4^+^ T cells, and particularly the PD-1^+^ CD4^+^ T cell subset, are crucial effectors that influence pembrolizumab efficacy in CTCL. In responders post-treatment, PD-1^+^ CD4^+^ T cells increase GZMB expression and move closer to tumor cells, similar to the granzyme-perforin-dependent tumor killing mechanism used by cytotoxic CD8^+^ T cells^45-48^. Previous studies have shown that antitumor activity is enhanced by a closer proximity of cytotoxic T cells to tumor cells^67,68^ and by removing inhibitory Tregs from the TME milleu^50^. The *SpatialScore* combines these principles by measuring the physical distances between PD-1^+^ CD4^+^ T cells and tumor cells (i.e., effector function) as well as PD-1^+^ CD4^+^ T cells and Tregs (i.e., suppressive function). The *SpatialScore* therefore reflects the balance of T cell effector activity versus suppression in the TME and the efficacy of PD-1 blockade in CTCL.

Third, pembrolizumab therapy leads to increased RNA and protein expression of CXCL13 in responder tumor cells (or kills cells with lower levels of expression), which may be advantageous in localizing effector PD-1^+^ CD4^+^ T cells within the TME. Overexpression of *CXCL13* is associated with improved clinical outcomes in breast^69^, colon^70^, ovarian^71^, lung^72^, and urothelial cancers^73^ as well as responsiveness to anti-PD-1 immunotherapy^72,74^. Additionally, upregulation of *CXCL13* can be accompanied by constitutive protein secretion^69,72^, which strongly attracts CXCR5^+^ CD4^+^ T cells^72^ and PD-1^+^ CD4^+^ T cells^75^ to the tumor site. A recent study also showed that the tumor-specific gene expression program of cytotoxic GZMB^+^ CD4^+^ T cells in bladder cancer treated with anti-PD-L1 therapy was marked by tumor overexpression of *CXCL13*^50^. Furthermore, CXCL13 null mice with bladder tumors did not respond to anti-PD-1 treatment and had a lower frequency of T cell infiltration compared their wild-type counterparts^73^. Collectively, these findings support a chemoattractant mechanism for the sustained clinical response to pembrolizumab therapy observed in responders: Upregulation of CXCL13 in tumor cells attracts effector PD-1^+^ CD4^+^ T cells to them, promoting successful antitumor activity. This aspect of pembrolizumab responsiveness is captured by the lower *SpatialScore* seen in responders and underscores the importance of T cell topography as a spatial biomarker.

There are important limitations to this study. Discovery of the *SpatialScore* was based on 14 patients. Larger studies are needed to determine if this spatial biomarker translates broadly for prediction of pembrolizumab response in CTCL and to establish a threshold value for the *SpatialScore* that can be used to stratify patients into probabilistic responders and non-responders. Additionally, larger studies might delineate more subtle features of the spatial architecture not detected in this study. Another limitation of this study is that the post-treatment tumor biopsies were collected at various time points, impairing our ability to directly assess modulation of T cell subsets at the precise time of therapeutic success or failure. Finally, our cohort is limited to advanced, heavily pre-treated CTCL patients. Further efforts should examine the therapeutic activity of PD-1 blockade in early stage and treatment naïve patients, since early tumors are generally more responsive to immunotherapy ^76-81^ and prior systemic treatment may alter the immune response to therapy. Despite these limitations, the robustness of our findings highlights that spatial organization and immune functionality are conserved in responders and non-responders, which reinforces the importance of these factors for determining pembrolizumab response in CTCL.

Future studies with scRNA-seq and paired T cell receptor sequencing are needed to better characterize the heterogeneity of PD-1^+^ CD4^+^ T cells across patient groups and to fully understand the cytotoxic potential of this crucial effector subset. Similarly, investigating the increased suppressive function of Tregs and the ICOS^+^ Treg subset in non-responders may reveal vulnerabilities that could be targeted in combination with PD-1 blockade. Given our findings that non-responders have an increased localization of Tregs and upregulation of *BRD3*, an epigenetic regulator, in their tumor cells pre-treatment, future studies should investigate the therapeutic potential of combining mogamulizumab (which depletes Tregs^82^) and/or bromodomain inhibitors (which inhibit BRD3 and decrease tumor cell proliferation^83^) with pembrolizumab therapy for CTCL. Further studies are also required to determine if CXCL13 expression levels—in both skin and blood—can serve as a surrogate of pembrolizumab activity and therefore guide ongoing treatment decisions. Such studies are especially important for advanced CTCL patients, where the burden of disease in the non-skin compartments often dictate clinical outcome.

This work provides an important conceptual foundation for improving the efficacy of anti-PD-1 immunotherapy in CTCL. We identify effector PD-1^+^ CD4^+^ T cells in responders and inhibitory ICOS^+^ Tregs in non-responders. The *SpatialScore* captures the interactions of these cell-types with tumor cells and reveals how PD-1 blockade manipulates the balance of effector and suppressive T cell function to achieve disease control. We anticipate that the cell-types comprising the *SpatialScore* will depend on the specific cancer and immunotherapy. The *SpatialScore* biomarker approach and the ability to measure it using a clinically accessible mIHC platform should enable its widespread use in cancer immunotherapy studies.

## Data Availability

All data generated or analyzed in this study are included in this Medrxiv article or available upon reasonable request.

## ACKNOWLEDGEMENTS

We thank all members of the Nolan laboratory for helpful discussions. We also thank Angelica Trejo, Pauline Chu, and Sarah Black (Departments of Microbiology & Immunology and Pathology) for excellent technical assistance, Sujay Vennam (Department of Pathology) for help with RNA-seq data processing, and Łukasz Kidziński (Department of Bioengineering) for statistical guidance. We are indebted to the patients and their families and to staff at all clinical sites. This work was supported by the National Institutes of Health (NIH) 2U19AI057229-16, 5P01HL10879707, 5R01GM10983604, 5R33CA18365403, 5U01AI101984-07, 5UH2AR06767604, 5R01CA19665703, 5U54CA20997103, 5F99CA212231-02, 1F32CA233203-01, 5U01AI140498-02, 1U54HG010426-01, 5U19AI100627-07, 1R01HL120724-01A1, R33CA183692, R01HL128173-04, 5P01AI131374-02, 5UG3DK114937-02, 1U19AI135976-01, IDIQ17X149, 1U2CCA233238-01, 1U2CCA233195-01 (G.P.N.), F32CA233203 (D.P.), T32AR007422 (D.P.), T32AI007290 (G.L.B.); The National Cancer Institute: U01CA154967 and UM1CA154967 (M.A.C., S.P.F.); Cancer Center Support Grant P30 CA015704. The Department of Defense (W81XWH-14-1-0180 and W81XWH-12-1-0591) (G.P.N.); The Food and Drug Administration (HHSF223201610018C and DSTL/AGR/00980/01) (G.P.N.); Cancer Research UK (C27165/A29073) (G.P.N.); The Bill and Melinda Gates Foundation (OPP1113682) (G.P.N.); The Cancer Research Institute (G.P.N.); The Parker Institute for Cancer Immunotherapy (G.P.N.); The Kenneth Rainin Foundation (2018-575) (G.P.N.); Celgene, Inc. (133826 and 134073) (G.P.N.); the Rachford & Carlotta A. Harris Endowed Chair (G.P.N.); and The Beckman Center for Molecular and Genetic Medicine (D.P., C.M.S., Y.H.K, G.P.N.). D.P. was also supported by Stanford Dean’s Fellowship and a Stanford Cancer Institute Fellowship. C.M.S. was supported by an Advanced Postdoc Mobility Fellowship from the Swiss National Science Foundation (P300PB_171189 and P400PM_183915) and an International Award for Research in Leukemia from the Lady Tata Memorial Trust, London, UK. S.S.B. was supported by a Bio-X Stanford Interdisciplinary Graduate Fellowship and Stanford Bioengineering. S.J. was supported by the Leukemia & Lymphoma Society Career Development Program and the Stanford Dean’s Fellowship.

## AUTHOR CONTRIBUTIONS

D.P. and C.M.S. conceived of the study. D.P., M.M., B.R.G., S.S.B., G.L.B., S.J., J.D., K.S.S., Y.G., and C.M.S. performed experiments and analyzed data. R.H.P, S.P.F., N.R., and M.A.C. provided clinical samples and created the tissue microarray. M.S.K. and Y.H.K coordinated trial protocols and provided clinical data. G.P.N. guided experiments and data analysis, with assistance from R.B.W. D.P. and G.P.N. wrote the manuscript, with input from all authors. All authors vouch for the accuracy of the data reported and approve the manuscript.

## METHODS

### Human subjects and clinical trial study design

The CITN-10 trial was a multicenter, phase II, single-arm clinical trial that investigated the efficacy of pembrolizumab in 24 patients with two common forms of relapsed/refractory CTCL, mycosis fungoides and Sézary syndrome^13^. Written informed consent was obtained from all enrolled patients. The use of their tissues for research was fully de-personalized and approved by the Stanford University IRB Administrative Panels on Human Subjects in Medical Research (HSR 46894). All patients had a clinicopathologically confirmed diagnosis of mycosis fungoides or Sézary syndrome (clinical stage IB to IV) that had relapsed, was refractory to, or had progressed after at least one standard systemic therapy. Exclusion criteria included central nervous system disease, active autoimmune disease, previous exposure to any anti-PD-1, anti-PD-L1, or anti-PD-L2 therapy, or treatment with radiotherapy or other anti-cancer agents within 15 weeks of the pre-treatment biopsy. Topical medications were not applied to the biopsied areas during treatment or within 8 weeks of the pre-treatment biopsy. Patients were treated with 2 mg/kg pembrolizumab by intravenous infusion every 3 weeks for up to 24 months^13^. Response and primary end point (overall response rate) were assessed by consensus global response criteria^23^.

### Sample collection and tissue microarray construction

Skin biopsy specimens were collected from the primary tumor site and FFPE histology blocks were generated according to standard pathology procedures. Pre-treatment biopsies were obtained prior to the first pembrolizumab infusion and post-treatment biopsies were collected during and at the conclusion of therapy (**Sup. Fig. 1, Sup. Table 1**). H&E-stained sections from all biopsies were reviewed by two board-certified pathologists (C.M.S. and R.H.P.). Fourteen of the 24 biopsy samples had adequate FFPE material to be assembled into a tissue microarray, and two to three cores of 0.6 mm diameter from the most infiltrated regions of each biopsy were digitally annotated and compiled into a tissue microarray. The tissue microarray was sectioned at 4-µm thickness and mounted onto Vectabond™-treated (Vector Labs, #SP-1800) square glass coverslips (22×22 mm, #1 1/2, Electron Microscopy Sciences, #72204-01).

### Immunohistochemistry

IHC for CD3 (clone CD3-12; Abd Serotec), CD4 (clone 4B12; Leica), CD8 (clone CD8/144B; Dako), FoxP3 (clone 236A/E7; Abcam), CD163 (clone 10D6; Thermo Fisher Scientific), PD-1 (clone NAT105; Cell Marque), PD-L1 (clone 22C3; Merck Research Laboratories), and PD-L2 (clone 3G2; Merck Research Laboratories) was performed as previously described^84^. Images were scored by CITN pathologists, and the positive percentage of the total mononuclear cell infiltrate was reported^13^.

### Multiplex immunohistochemistry and analysis

mIHC was performed as previously described^85^. Briefly, 4-µm FFPE tissue sections were baked for 1 h at 60 °C, dewaxed, and stained on a BOND Rx autostainer (Leica) according to Opal Multiplex IHC assay (Akoya Biosciences)s protocol with the following changes: additional high stringency washes were performed after the secondary antibody and Opal fluor applications using high-salt TBST (0.05M Tris, 0.3M NaCl and 0.1% Tween-20, pH 7.2-7.6), and TCT was used as the blocking buffer (0.05M Tris, 0.15M NaCl, 0.25% Casein, 0.1% Tween 20, pH 7.6 +/- 0.1). The antibody panel was stained in the following order, with antibody stripping between positions. Each primary antibody was incubated for 60 min, followed by 10-min incubation with secondary antibody (OPAL polymer HRP mouse plus rabbit, Akoya Biosciences), followed by application of the tertiary TSA-amplification reagent (OPAL Fluor, Akoya Biosciences) for 10 min. Positions were as follows: position 1: CD8 (clone CD8/144B, DAKO #M7103; concentration 0.8 µg/ml; OPAL Fluor 520); position 2: CD25 (clone 4C9, Cell Marque #125M-14; concentration 0.17 µg/ml; OPAL Fluor 540); position 3: CD3 (clone SP7, Thermo Fisher Scientific #RM-9107; concentration 0.06 µg/ml; OPAL Fluor 570); position 4: PD-1 (clone EPR4877(2), Abcam #ab137132, concentration 1.0 µg/ml; OPAL Fluor 650); position 5: CD7 (clone MRQ-56; Cell Marque #107M-24; concentration 1.18 µg/ml; OPAL Fluor 690); position 6: CD4 (clone EP204, Epitomics; #AC0173A; concentration 0.08 µg/ml; OPAL Fluor 480); and position 7: FoxP3 (clone 236A/E7; eBioscience #14-4777-82; 5.0 µg/ml; OPAL Fluor 620). Subsequently, slides were stained with Spectral DAPI (Akoya Biosciences) for 5 min, rinsed, and mounted with Prolong Gold Antifade reagent (Thermo Fisher Scientific #P36930). After curing for 24 h at room temperature in the dark, images were acquired on a Vectra Polaris automated quantitative pathology imaging system (Akoya Biosciences). The raw images were spectrally unmixed using the Phenoptics inForm software (Akoya Biosciences) and exported as multi-image TIFF files.

After fluorescent imaging, the slides were de-coverslipped, loaded onto the BOND Rx autostainer, stripped of bound antibody, and a post-mIHC staining for CXCL13 (goat polyclonal, R&D Systems #AF801; concentration 0.5 µg/ml; incubation 60 min) was performed. Bound antibody was revealed by anti-goat HRP secondary ImmPress HRP (Vector Labs #MP-7405; incubation 12 min), followed by DAB chromogen using the BOND Polymer Refine Detection kit (Leica) according to the manufacturer’s instructions. After counterstaining with hematoxylin, slides were dry-mounted and scanned on an Aperio AT turbo digital slide scanning system (Leica).

HALO software (Indica Labs) was used to perform single-cell analysis of mIHC images. Cells were visualized based on nuclear and cytoplasmic stains, and mean pixel fluorescence intensity in the applicable compartments of each cell were measured (i.e., CD4 in the cytoplasmic compartment and FoxP3 in the nuclear compartment). A mean intensity threshold above background was used to determine positivity for each fluorochrome, thereby defining cells as either positive or negative for each marker. The data was then used to define co-localized populations, including PD-1^+^ CD4^+^ T cells, tumor cells, and Tregs. Spatial positions were extracted for each cell, and the spatial distances and ratios between these three cells types were calculated as detailed below. Performance of the *SpatialScore* biomarker was evaluated with the easyROC interface available with the R package shiny^86^. CXCL13 IHC images were scored using a classifier method for the DAB stain based on optical density to obtain the positive percentage of the total mononuclear cell infiltrate per spot.

### CODEX antibodies

For CODEX, purified, carrier-free monoclonal and polyclonal anti-human antibodies were purchased from commercial vendors (**Sup. Table 1**). Conjugations to maleimide-modified short DNA oligonucleotides (TriLink) were performed at a 2:1 weight/weight ratio of oligonucleotide to antibody, with at least 100 µg of antibody per reaction, as previously described^19^. Conjugated antibodies were validated and titrated under the supervision of a board-certified pathologist (C.M.S.).

### CODEX multiplex tissue staining and imaging

The CODEX experiment was performed as previously described^19^. Briefly, the coverslip was deparaffinized and rehydrated, and heat-induced epitope retrieval was performed using Dako target retrieval solution, pH 9 (Agilent, #S236784-2) at 97 °C for 10 min. The coverslip was stained with an antibody cocktail (**Sup. Table 2**) to a volume of 100 µl overnight at 4 °C in a sealed humidity chamber on a shaker. After multiple fixation steps using 1.6% paraformaldehyde, 100% methanol, and BS3 (Thermo Fisher Scientific, #21580), the coverslip was mounted onto a custom-made acrylic plate (Bayview Plastic Solutions). Imaging was performed with a Keyence BZ-X710 inverted fluorescence microscope equipped with a CFI Plan Apo λ 20x/0.75 objective (Nikon), an Akoya CODEX microfluidics instrument, and CODEX driver software (Akoya Biosciences). Light exposure times and the arrangement of cycles are outlined in **Sup. Table 2**. At the conclusion of the CODEX multicycle reaction, H&E staining was performed, and images were acquired in brightfield mode.

### Data processing of CODEX images

Raw TIFF image files were processed using the CODEX Toolkit as previously described. After processing, the staining quality for each antibody was visually assessed in each tissue microarray spot, and cell segmentation was performed using the DRAQ5 nuclear stain. Marker expression was quantified, and single-cell data were saved as FCS files, which were then imported into CellEngine (https://cellengine.com) for cleanup gating. This resulted in a total of 117,170 cells across all tissue microarray spots.

After cleanup gating, FCS files were exported from CellEngine, imported into VorteX clustering software^87^, and subjected to unsupervised X-shift clustering using an angular distance algorithm. Clustering was based on all antibody markers except CD11b, CD16, CD164, CCR4, CCR6, EGFR, and p53. The optimal cluster number was guided by the elbow point validation tool in VorteX, resulting in 78 clusters. Clusters were manually verified and assigned to cell-types based on morphology in H&E and fluorescent CODEX images and on their marker expression profiles. Clusters with similar features were merged, resulting in 21 cell-type clusters. The expression frequencies of ICOS, IDO, Ki-67, and PD-1 were determined for the T cell and tumor cell clusters by manual gating in CellEngine for each tissue microarray spot, with visual comparison to the raw fluorescent image.

### Cellular neighborhood identification

CN identification was performed using a custom *k*-nearest neighbors’ algorithm in Python^19^. For each of the 117,220 cells in this experiment, the window size was set at 10, capturing the center cell and its nine nearest neighboring cells, as measured by the Euclidean distance between X/Y coordinates. To identify 10 CNs, these windows were then clustered by the composition of their microenvironment with respect to the 21 cell-types that were previously identified. This resulted in a vector for each window containing the frequency of each of the 21 cell-types amongst the 10 neighborhoods. These windows were then clustered using Python’s *scikit-learn* implementation of MiniBatchKMeans with *k*=10. Each cell was then allocated to the same CN as the window in which it was centered. All CN assignments were validated by overlaying them on the original fluorescent and H&E-stained images.

### Calculation of spatial distances and ratios between cell-types

The X/Y coordinates for each cell type were determined during cellular segmentation, as described above. The minimal distance between each cell-type and its nearest other cell-types, and the averages of these minimal distances per tissue spot, were calculated in R. Given our interest in the relationship of cell distances between three cell-types (i.e., effector T cells (CT1), tumor cells (CT2) and Tregs (CT3)), we calculated the ratio of the minimal distances between CT1—CT2 (right distance) versus CT1—CT3 (left distance). This distance ratio represented the *SpatialScore*. To assess whether these ratios were significantly different from those of a random sample, we performed the following analysis per spot: For the number of CT1 cells in each spot, we randomly selected the same number of non-CT1 (nCT1) cells. For each of these nCT1 cells, we calculated the ratio of the minimal distances (nCT1—CT2 / nCT1—CT3) and determined the mean of this sample. We repeated this random sampling 100 times, and the average of all the means was reported. Distribution of the random values was assessed by the quant output variable, which indicates how many of the random means are smaller than the measured means. For instance, a quant of 97 indicates that 97% of the random means are smaller than the measured means. Thus, quant values closer to 100 or 0 indicate that the measured means are not random (**Sup. Fig. 4j-m**).

### Laser-capture microdissection (LCM)

Two serial sections of the tissue microarray were taken at 7 µm thickness and mounted onto frame slides with polyethylene naphthalate membranes (Thermo Fisher Scientific, #LCM0521). Slides were immersed for 20 s each in xylene (three times), 100% ethanol (three times), 95% ethanol (two times), 70% ethanol (two times), water, hematoxylin (Dako, #S3309), water, bluing reagent (Thermo Fisher Scientific, #7301), water, 70% ethanol (two times), 95% ethanol (two times), 100% ethanol (three times), and xylene (three times). Immediately after staining, cells were dissected from every tissue microarray spot on an ArcturusXT LCM System (Thermo Fisher Scientific) using the ultraviolet laser to cut out the desired region and the infrared laser to adhere the membrane to a CapSure HS LCM Cap (Thermo Fisher Scientific, #LCM0215). A tissue area containing roughly 1000 mononuclear cells was captured from each spot, with cell numbers determined based on density estimates by cell counting in an adjacent H&E-stained section. If a core had more than 1000 mononuclear cells, a tissue fragment containing around 1000 mononuclear cells was dissected from that core. If a core had less than 1000 cells, tissue fragments from corresponding cores on the serial section membrane were combined in the same LCM cap to obtain approximately 1000 cells. After microdissection, the caps were sealed using 0.5-ml tubes (Thermo Fisher Scientific, #N8010611) and stored at −80 °C until cDNA library preparation.

### Preparation of cDNA libraries and RNA sequencing

Sequencing libraries were prepared according to the Smart-3Seq protocol for LCM HS caps, as previously described with slight modifications^21^. Briefly, 10 µl of lysis mix consisting of 40% (v/v) 5 M trimethylglycine solution (Sigma, #B0300), 20% (v/v) 10 mM nuclease-free dNTP mix (Thermo Fisher Scientific, #R0192), 10% (v/v) 20 µM first-strand primer in TE buffer (1S, /5Biosg/GT GAC TGG AGT TCA GAC GTG TGC TCT TCC GAT CTT TTT TTT TTT TTT TTT TTT TTT TTT TTT TV; Integrated DNA Technologies), 10% (v/v) Triton-X 100 (Sigma #T8787; diluted to 0.5% v/v in molecular biology-grade water), and 20% (v/v) Proteinase K (New England Biolabs, #P8107S; diluted to 0.125 mg/ml in in molecular biology-grade water) was added to the center of each LCM cap. Caps were sealed with 0.2 ml low-retention PCR tubes (Corning, #PCR-02-L-C) and incubated on a pre-warmed metal CapSure incubation block (Thermo Fisher Scientific, #LCM0505) at 60 °C in an incubator. Then, tubes were briefly centrifuged, and 10 µl of template-switching reverse-transcription (TS-RT) FFPE LCM mix consisting of 40% (v/v) 5x SMARTScribe first-strand reaction buffer (Clontech, #639537), 20% (v/v) 20 mM DTT (Clontech, #639537), 10% (v/v) 20x RNase inhibitor (Thermo Fisher Scientific, #AM2694), 4% (v/v) 50 µM second-strand primer in TE buffer (2S, /5Biosg/CT ACA CGA CGC TCT TCC GAT CTN NNN NrGrG rG; Integrated DNA Technologies), 4% (v/v) 200 mM MgCl2 (Sigma, #63069), 2% 5 mM proteinase K inhibitor (EMD Millipore, #539470), and 20% (v/v) 100 U/µl SMARTScribe reverse transcriptase (Clontech, #639537) was added. Samples were incubated in a programmable thermal cycler (42 °C for 30 min, 70 °C for 10 min, 4 °C hold), and 1.25 µl of a unique P5 primer and 1.25 µl of a universal P7 primer (2 µM in TE buffer each; Integrated DNA Technologies; sequences available upon request) and HiFi HotStart ReadyMix (Kapa, #KK2601) were then added, followed by 22 cycles of PCR amplification (98 °C for 45 s; 22 cycles at 98 °C for 15 s, 60 °C for 30 s, 72 °C for 10 s; then 72 °C for 60 s, and 4 °C hold). Amplified cDNA was next purified with SPRI bead mix (Beckman Coulter, #B23317) and a magnetic separation block (V&P Scientific, #VP772F4). Finally, the samples were washed with 80% ethanol and resuspended in TE buffer to yield the sequencing-ready library.

Libraries were profiled for size distribution on an Agilent 2200 TapeStation with High Sensitivity D1000 reagent kits and quantified by qPCR with a dual-labeled probe as previously described^88^. Libraries were excluded if <40% of their transcripts were within a 165-500 bp range. A total of 64 libraries were mixed to equimolarity, according to the qPCR measurements. The RNA libraries were sequenced on an Illumina NextSeq 500 instrument with a High Output v2.5 reagent kit (Illumina, #20024906) to a minimum sequencing depth of1.5 M reads per sample (mean: 3.7 M) and minimum uniquely aligned reads of 364,468 per sample (mean: 916,607) using read lengths of 76 nucleotides (nt) for read 1 and 8 nt for read 2. On average, we obtained reads from 11,166 genes per sample (median: 11,267) and 379,615 unique transcripts per sample (median: 336,005), which is comparable to previously published FFPE-based RNA-seq studies of human cancers^21^.

### Processing of RNA-seq data

Base calls from the NextSeq were de-multiplexed and converted to FASTQ format with bcl2fastq (Illumina). The five-base unique molecular identifier (UMI) sequence and the G-overhang were extracted from FASTQ data, and A-tails were removed with umi_homopolymer.py (github.com/jwfoley/3SEQtools). Reads were aligned and further processed to remove duplicates using STAR (github.com/alexdobin/STAR). Bulk gene expression profiles were transcript per million (TPM) normalized and log2 transformed. Differences in *CXCL13* and *CXCR5* expression between groups were modeled with Linear Mixed Effects Models on a per spot basis using the lmer function from package lme4 (v1.1.21)^89^ and taking the patient intercept as a random effect. The pairwise p-values were derived from t-ratio statistics in the contrast analysis using the lmerTest (v3.1.2)^90^ and corrected for multiple hypothesis testing using the Holm Bonferroni method implemented in the modelbased (v0.1.2) package (github.com/easystats/modelbased).

### Principal component analysis (PCA) immune scores

PCA scores and principal component 1 (PC1) coefficients were computed for the normalized bulk RNA-seq data on a per spot basis using the prcomp function in base R. The IFN-γ score was calculated using the six gene signature published by Ayers et al.^39^. The TGF-β score was calculated using the 15 gene signature published by Mariathasan et al.^40^. The immune activation and immunosuppression scores were computed using the genes listed in **Fig. 2g**. Differences in PC1 scores between patient groups were modeled using Linear Mixed Effects Models on a per spot basis using the lmer function from package lme4 (v1.1.21)^89^ and taking the patient intercept as a random effect. Differences between responders and non-responders as well as pre-treatment and post-treatment samples were modeled as fixed effects and tested using Satterthwaite’s degrees of freedom method. The pairwise *p* values were derived from t-ratio statistics in the contrast analysis using the lmerTest (v3.1.2)^90^ and corrected for multiple hypothesis testing using the Holm Bonferroni method implemented in the modelbased (v0.1.2) package (github.com/easystats/modelbased).

### Identifying bulk RNAseq gene signatures associated with tumor cells and the *SpatialScore*

LASSO regression models were used to find genes predictive of tumor cells and the *SpatialScore*. These models were estimated using the LassoCV object in the scikit-learn python package. Six-fold cross validation was used to select the optimal regularization parameter. Specifically, an L1-regularized linear model was fit to predict the frequency of tumor cells from the gene expression data per tissue microarray spot. For this model, the response variable was the log transformed per spot percentage of tumor cells. A pseudo-count of 1% was added to genes. The features utilized as predictors were the per spot log transformed TPM counts and the log frequency of CD4^+^ T cells. Genes with positive nonzero coefficients were interpreted as positively associated with tumor cells. Similarly, an L1-regularized linear model was fit to predict the *SpatialScore* from the gene expression data on a per spot basis. For this model, the response variable was the log transformed *SpatialScore* distance ratio. The features used as predictors were the per spot log transformed TPM counts. Genes with nonzero coefficients were selected in figures as predictive.

### CIBERSORTx signature matrix

To generate a CSx signature matrix, we used a publicly available scRNA-seq dataset from Gaydosik et al.^59^ that was obtained from skin biopsies of five CTCL patients. Datasets were downloaded from the Gene Expression Omnibus (GEO) database (accession code GSE128531), and single-cell profiles were combined and analyzed using the Seurat R package (3.1.4)^91^. Cells with between 500 and 7500 genes detected and less than 10% mitochondrial transcripts were included in the analysis.

Data were log10normalized and clustered with the Louvain method^92^ based on the first 13 PCs and resolution of 1.8. Cell clusters were visualized using Uniform Manifold Approximation and Projection (UMAP)^93^, with the same PCs. Major cell-types were assigned according to expression of corresponding marker genes (**Sup. Fig. 6a, Sup. Table 4**). Fibroblasts and pericytes were merged into a stromal cluster. The T cell cluster was divided into CD4^+^ T cells, CD8^+^ T cells, Tregs, and tumor cells based on the expression of certain T cell and tumor marker genes (**Sup. Fig 6b, Sup. Table 4**). Tumor cells from patient CTCL-5 were excluded due to extreme heterogeneity. T cell clustering was based on the first 15 PCs and resolution of 1.9. The same PCs were used to generate UMAP projections for the T cell clusters. A matrix of single cells and their assigned cell-type identities was used to create a signature matrix using the CSx (v.1.0) website^22^ (code available from https://cibersortx.stanford.edu/). There was good correlation between the CSx and CODEX cell-type clusters (**Sup. Fig 5c**).

### CIBERSORTx deconvolution

The signature matrix was used to deconvolve tumor cell gene expression in CSx (v.1.0) with the CSx website (arguments used: rmbatchSmode = T, QN = F)^22^. Log2 fold changes were computed for every deconvolved gene across patient groups. Differences in gene expression between patient groups were modeled with Linear Mixed Effects Models on a per spot basis using the lmer function from package lme4 (v1.1.21)^89^ and taking the patient intercept as a random effect. The *p* values were derived using Satterthwaite’s degrees of freedom method, implemented in the lmerTest (v3.1.2)^90^ package. The *p* values were adjusted with the Benjamini-Hochberg correction using the p.adjust function in R. Volcano plots were generated using the ggplot2 (3.3.0)^94^ and ggrepel (0.8.1)^95^ packages in R. Genes with Benjamini-Hochberg-adjusted *p* < 0.1 were considered significant (**Fig. 5i-j, Sup. Fig. 5d**).

CSx-deconvolved *CXCL13* expression in tumor cells was log2 transformed on a per spot basis. Differences in *CXCL13* expression between patient groups were modeled using Linear Mixed Effects Models on a per spot basis using the lmer function from package lme4 (v1.1.21) and taking the patient intercept as a random effect. The pairwise *p* values were derived from t-ratio statistics in the contrast analysis using the lmerTest (v3.1.2) and corrected for multiple hypothesis testing using the Holm Bonferroni method implemented in the modelbased (v0.1.2) package (github.com/easystats/modelbased). To examine *CXCL13* expression in tumor cells on a per patient basis, the mean from biological replicates was computed before plotting the log2 normalized CSx-deconvolved *CXCL13* expression. The Wilcoxon signed-rank test was used to evaluate whether patient-matched CSx-deconvolved CXCL13 expression in the tumor cells was different between groups.

### Statistical analysis

Statistical analyses were performed with R and Prism v8 (GraphPad Software, Inc). Results with *p* < 0.05 were considered significant, unless otherwise stated. For pre-treatment differences between responders and non-responders, the significance was tested using a two-sided Wilcoxon’s rank-sum test. For differences across patient groups (i.e., responders and non-responders pre-treatment, responders pre- and post-treatment, non-responders pre- and post-treatment, and responders and non-responders post-treatment), the significance was tested using a linear mixed-effect model with Bonferroni’s corrections for multiple comparisons. Pre-to post-treatment pairwise statistical significance for individual patients was tested using a two-sided Wilcoxon’s signed-rank test. Correlations were evaluated with the non-parametric Spearman test. The investigators were not blinded to allocation during experiments and outcome assessment. No sample-size estimates were performed to ensure adequate power to detect a pre-specific effect size. \**p* < 0.05, \*\**p* < 0.01, \*\*\**p* < 0.001, \*\*\*\**p* < 0.0001.

## SUPPLEMENTARY FIGURE AND TABLE LEGENDS

**Supplementary Fig. 1.**
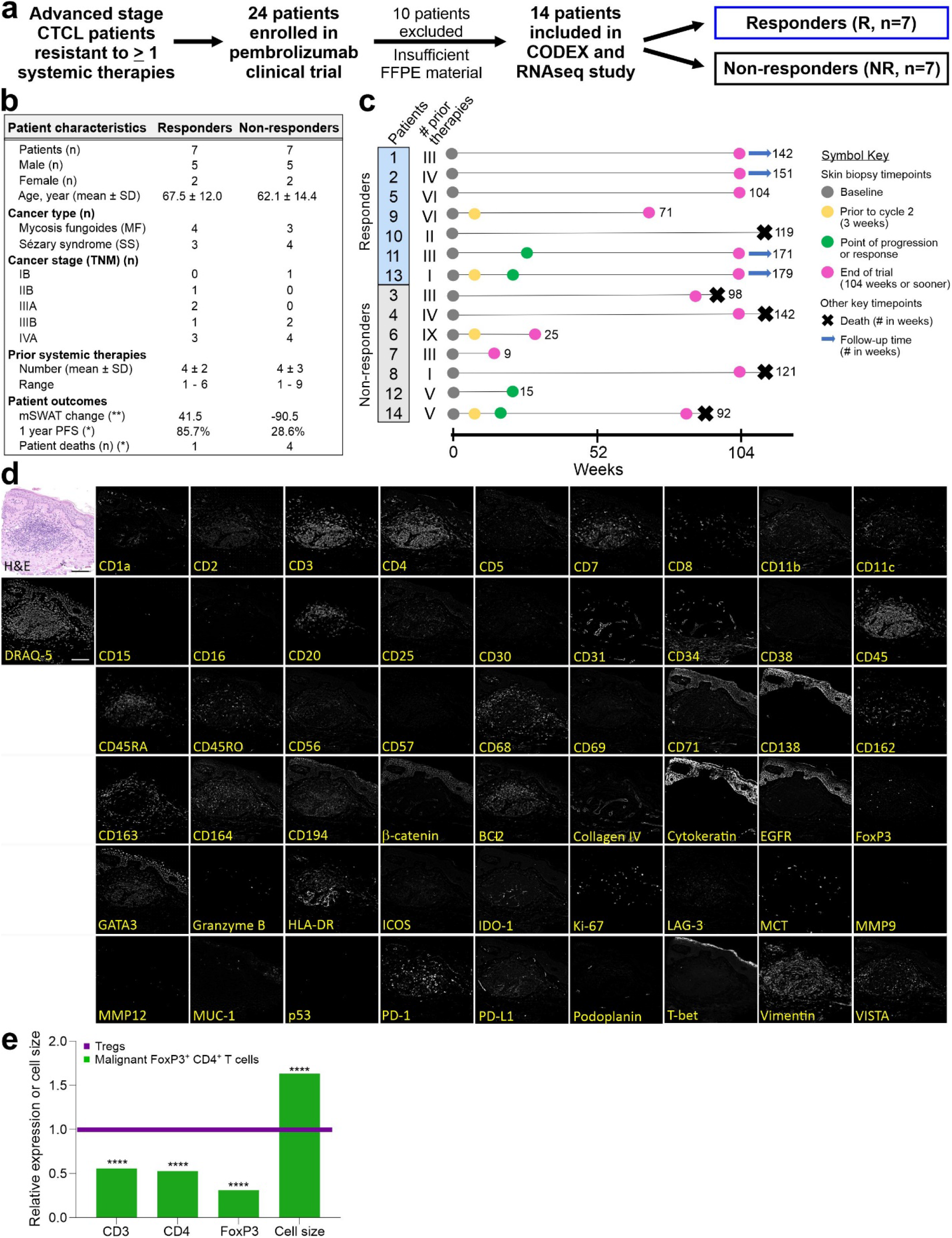
Study design and CODEX experimental validation. **a**, Framework for the 14 included CTCL patient samples used for CODEX tissue imaging, RNA-seq and mIHC imaging. **b**, Patient characteristics for the 7 responders and 7 non-responders. Significant differences between responders and non-responders were only noted for patient outcomes: the change from baseline in skin by the modified Severity Weighted Assessment Tool (mSWAT) (**, *p*=0.0019 by Wilcoxon’s rank sum test), the 1 year progression free survival (PFS) (*, *p*=0.0306 by Wilcoxon’s rank sum test), and patients deaths (*, *p*=0.0168 by log rank test in Kaplan-Meier overall survival curve; see **Fig. 1b**). **c**, Swimmer plot of individual patients, depicting treatment history and timepoints in weeks for every tumor biopsy used in this study. Lines represent overall survival in weeks. A rightward arrow indicates that the patient was still alive following the final tumor collection. A rightward X indicates that the patient died. **d**, Validation of the 55-marker CODEX antibody panel used to stain the CTCL skin tissue microarray. Images of a single microarray spot are depicted in false gray color for each marker. H&E staining is also shown. Scale bars, 100 µm. **e**, Average expression of CD3, CD4, FoxP3, and cell size (measured in pixels/cell) of malignant FoxP3^+^ CD4^+^ T cells from responder patient 2 (green bars), relative to Tregs from all patients (purple line). *P* values were calculated by two-sided Wilcoxon’s rank-sum tests (****, *p*<0.0001).

**Supplementary Fig. 2.**
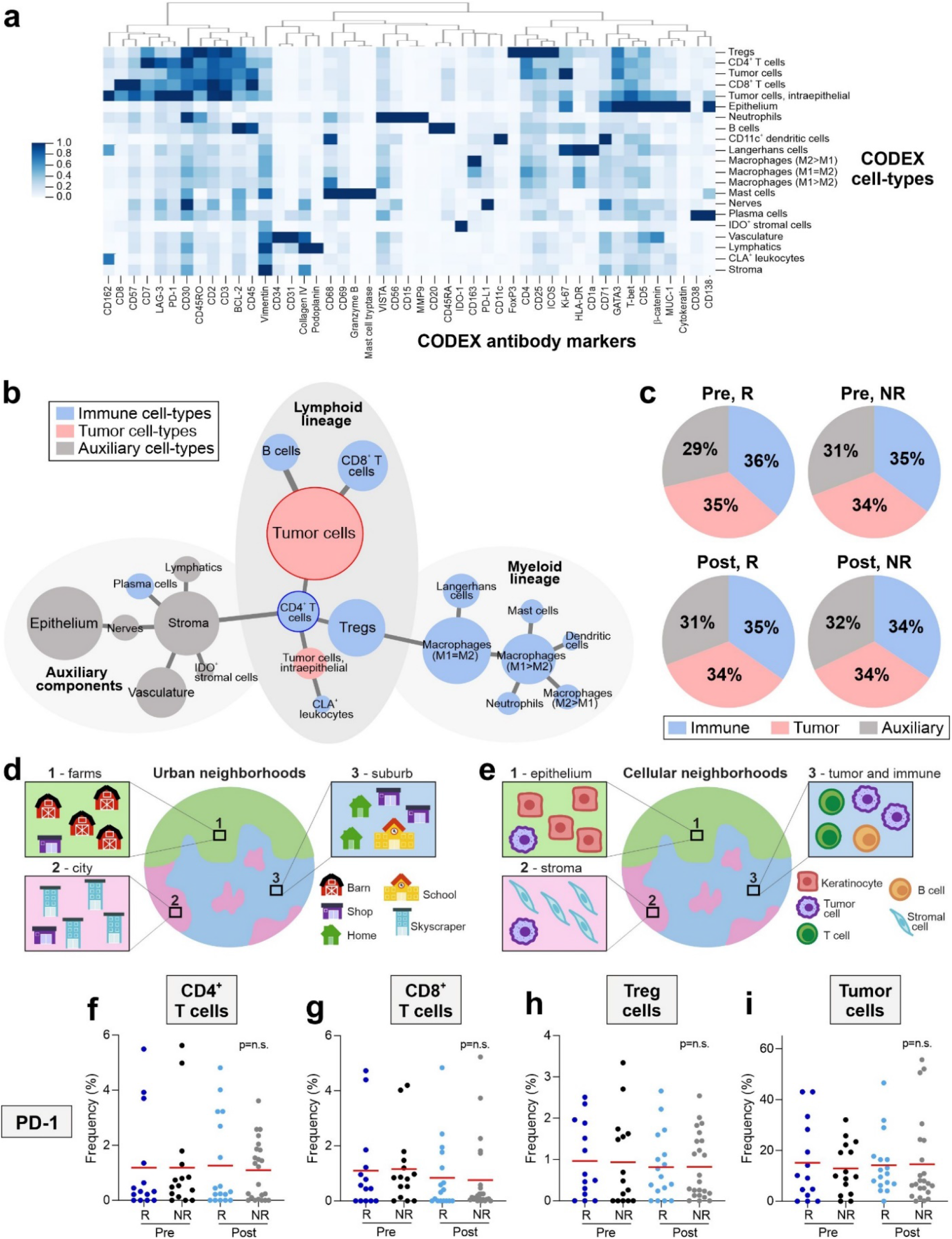
Characterizing CODEX cell-types, the cellular neighborhood concept, and PD-1 positivity. **a**, Heatmap of CODEX antibody marker expression in each of the 21 identified cell-types. **b**, Minimal spanning tree of the 21 cell-types, which are colored blue (immune cell-types), red (tumor cell-types) and gray (auxiliary cell-types). **c**, Frequencies of tumor, immune and auxiliary cell-types for each patient group (p=n.s. for all comparisons). **d-e**, Conceptual neighborhood schematic showing how urban neighborhoods are determined based on their composition of buildings (**d**) and cellular neighborhoods based on cell-types (**e**). **f-i**, Frequencies of PD-1^+^ CD4^+^ T cells (**f**), PD-1^+^ CD8^+^ T cells (**g**), PD-1^+^ Tregs (**h**), and, PD-1^+^ tumor cells (**i**) per tissue microarray spot in responders and non-responders pre- and post-treatment (mean, red bar). *P* values were calculated with a linear mixed-effect model with Bonferroni’s corrections for multiple comparisons.

**Supplementary Fig. 3.**
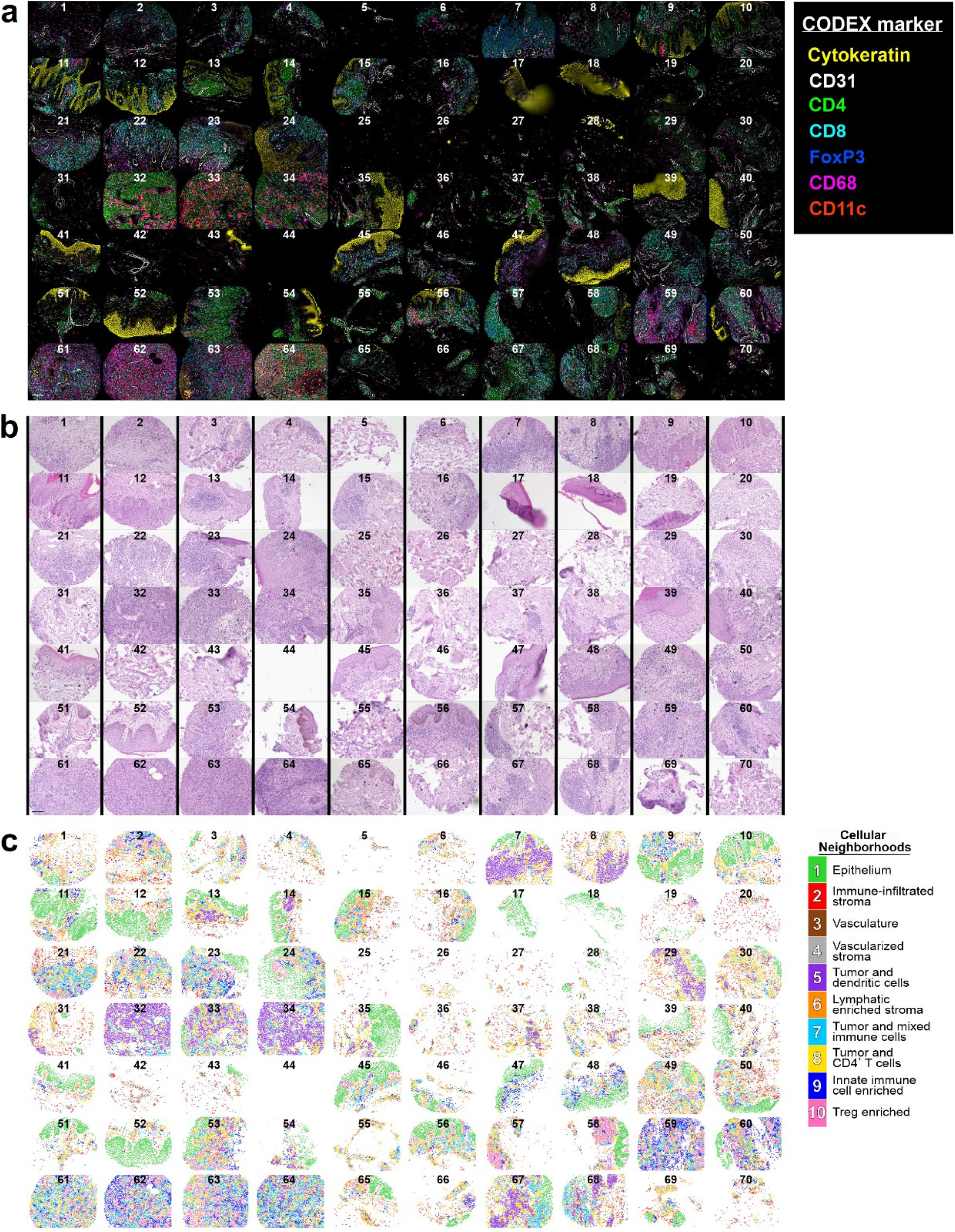
CODEX, H&E, and cellular neighborhood images for the CTCL tissue microarray. **a**, CODEX seven-color overlay image of the CTCL tissue microarray. Scale bar, 100 µm. **b**, Corresponding H&E image of the CTCL tissue microarray. Scale bar, 100 µm. **c**, Dot plot Voronoi diagram of the 10 identified CNs, colored according to the corresponding legend.

**Supplementary Fig. 4.**
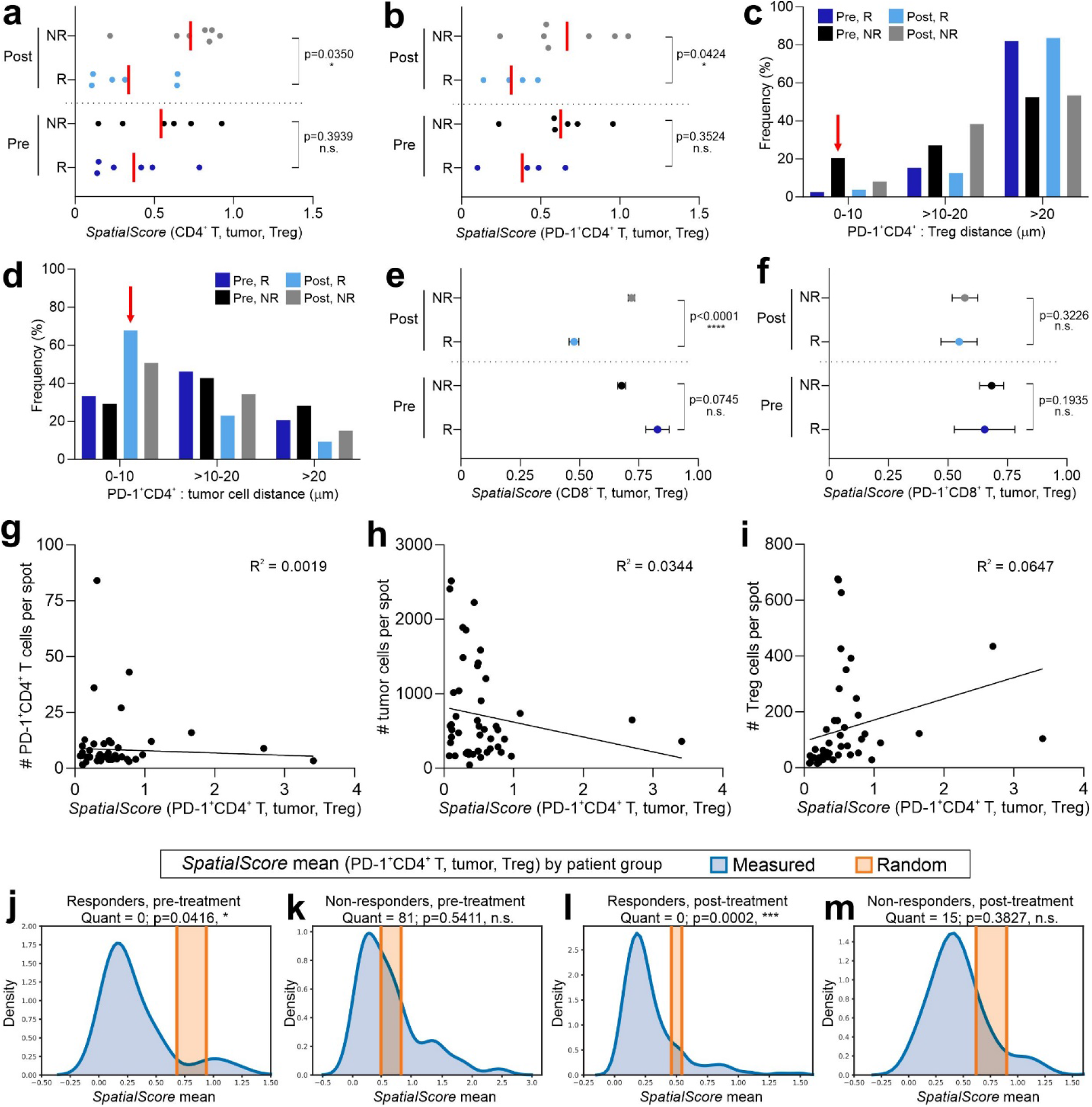
Detailing the *SpatialScore* in CTCL. **a-b**, *SpatialScore* based on all CD4^+^ T cells (**a**) and PD-1^+^ CD4^+^ T cells (**b**) on a per patient basis across patient groups (mean, red bar). Patients were excluded from this analysis if they did not have at least 10 CD4^+^ or PD-1^+^ CD4^+^ T cells. *P* values were calculated by two-sided Wilcoxon’s rank-sum tests, with no adjustments for multiple comparisons. **c**, Frequency distribution of the physical distances in the tissue between PD-1^+^ CD4^+^ and Tregs by patient group, with the red arrow showing that these two cell-types are closest in non-responders pre-treatment. **d**, Frequency distribution of the distances between PD-1^+^ CD4^+^ and tumor cells by patient group, with the red arrow showing that these two cell-types are closest in responders post-treatment. **e-f**, *SpatialScore* on a per cell basis using CD8^+^ T cells (**e**) and PD-1^+^ CD8^+^ T cells (**f**) for each patient group (mean ± s.e.m.). *P* values were calculated by two-sided Wilcoxon’s rank-sum tests, with no adjustments for multiple comparisons. **g-i**, Correlations of the number of PD-1^+^ CD4^+^ T cells (**g**) tumor cells (**h**), and Tregs (**i**) relative to the *SpatialScore* per tissue microarray spot. Correlations were evaluated with the Spearman test. **j-m**, Density plots of the measured mean *SpatialScore* distribution (blue) and corresponding random mean with its standard deviation (orange) for responders pre-treatment (j) non-responders pre-treatment (k) responders post-treatment (l), and non-responders post-treatment (m). *P* values were calculated by two-sided Wilcoxon’s rank-sum tests, with no adjustments for multiple comparisons. The Quant values correspond to the percentage of randomly measured values that are smaller than the measured value; values closer to 0 or 100 indicate that the measurement is not random.

**Supplementary Fig. 5.**
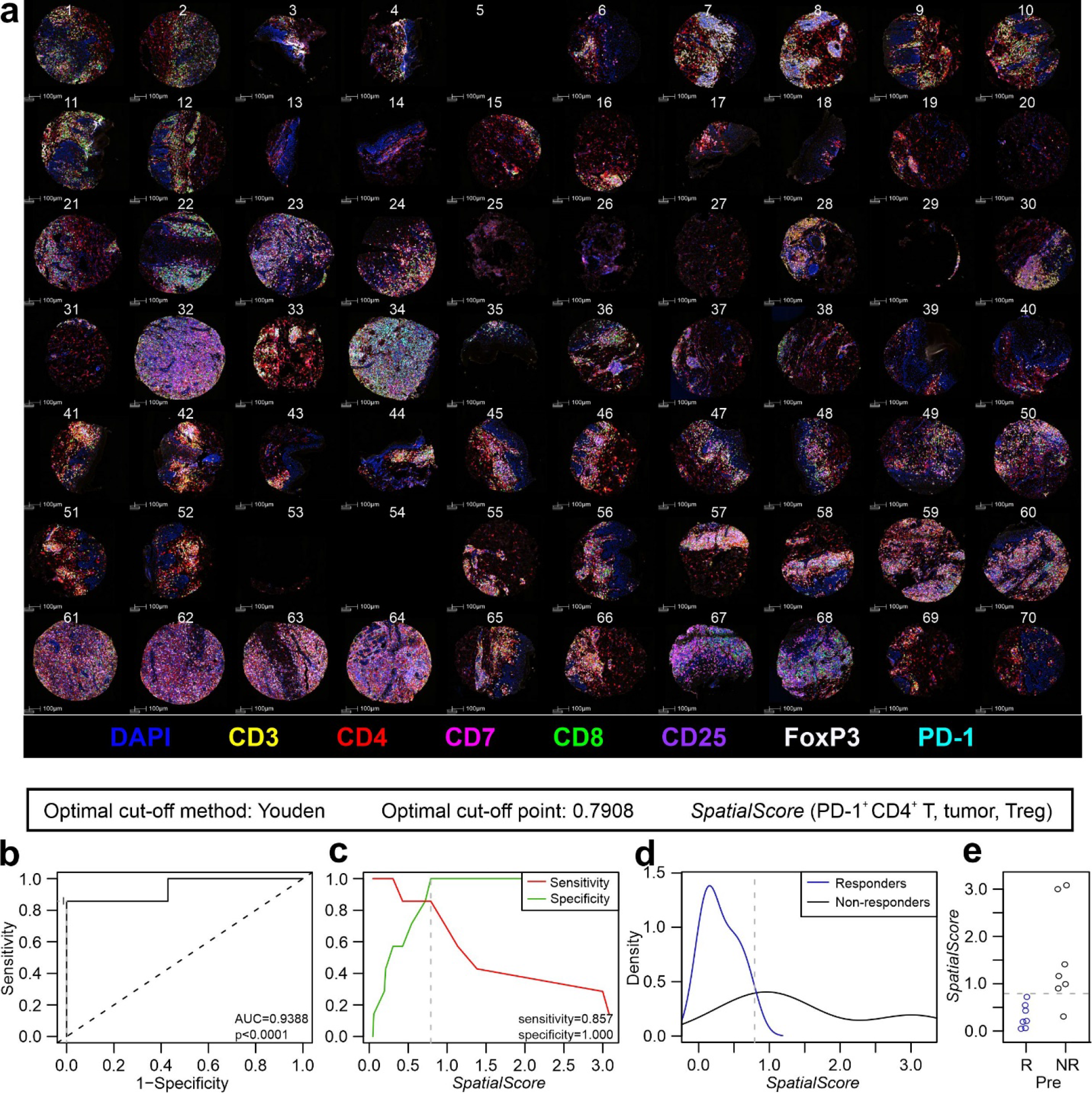
Validation of the *SpatialScore* biomarker using a clinically accessible mIHC platform. **a**, Vectra mIHC staining of the CTCL tissue microarray with an eight-color overlay image, including DAPI (blue), CD3 (yellow), CD4 (red), CD7 (magenta), CD8 (green), CD25 (purple), FoxP3 (white), and PD-1 (cyan). Scale bars, 100 µm. **b-e**, Biomarker performance measures, using an optimal cut-off point of 0.7908, for the pre-treatment *SpatialScore* calculated from data obtained with the Vectra mIHC platform including an ROC curve (**b**),sensitivity and specificity plots (**c**), density plots (**d**), and patient-based scatter plots (**e**), whereby 100% of responders were below the *SpatialScore* cut-off point and 85.7% of non-responders were above the cut-off point).

**Supplementary Fig. 6.**
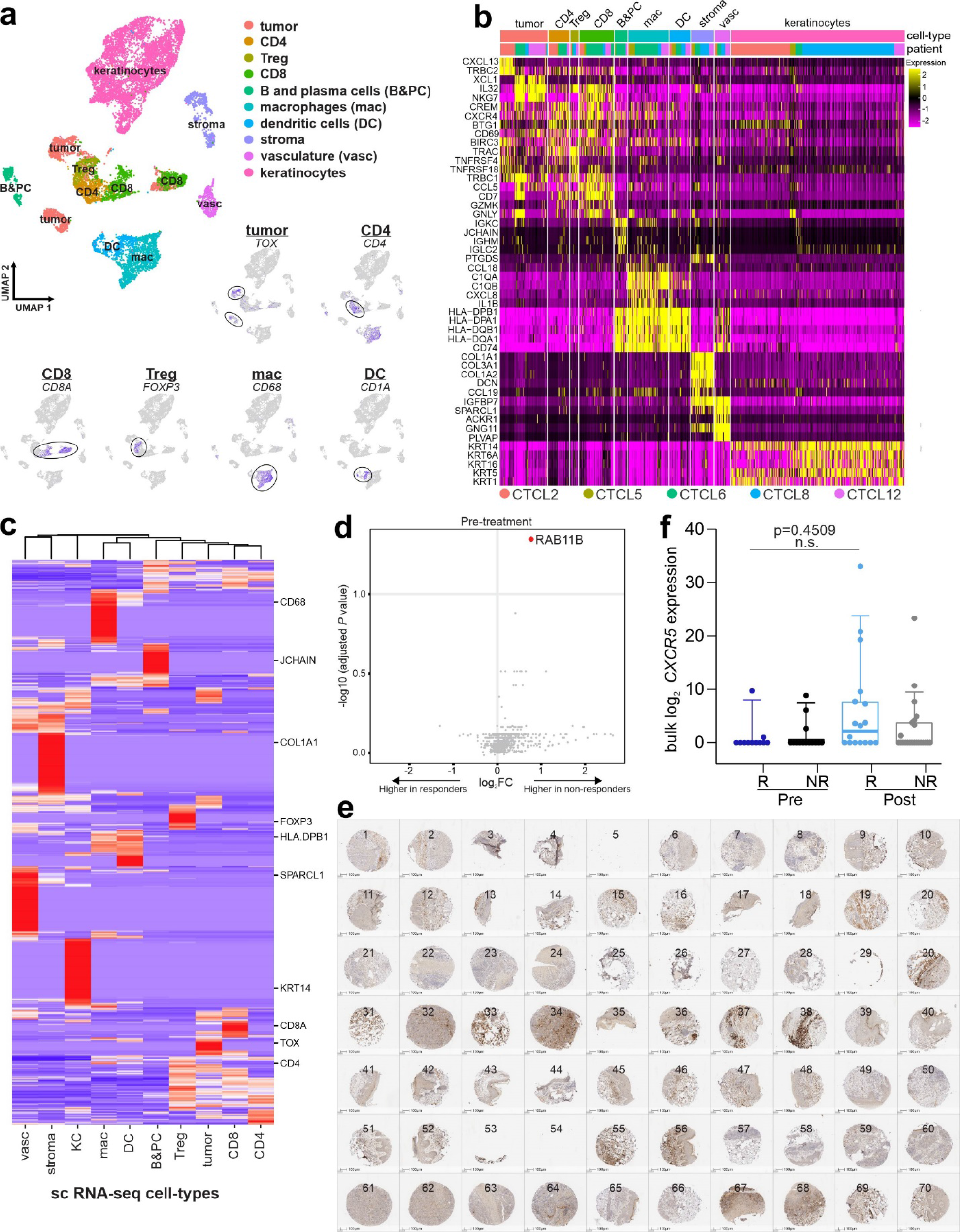
Characterizing CIBERSORTx cell-types and CXCL13 expression patterns. **a**, UMAP of 10 major cell-types identified in the Gaydosik et al.^59^ CTCL scRNA-seq dataset (see **Methods**), with feature plots showing expression for a subset of marker genes including *TOX* (tumor cells), *CD4* (CD4^+^ T cells), *CD8A* (CD8^+^ T cells), *FOXP3* (Tregs), *CD68* (macrophages), and *CD1A* (dendritic cells). **b**, Heatmap of the top differentially expressed genes (rows) for the 10 cell-types, corresponding to the top color bar. The other color bar corresponds to patient samples from the Gaydosik et al.^59^ CTCL scRNA-seq dataset (legend is at the bottom of the heatmap). **c**, Signature matrix, highlighting key marker selected genes (rows), used to enumerate cell-type fractions and resolve gene expression profiles from CTCL bulk RNA-seq. **d**, Differential expression of CSx-resolved tumor cell genes in responders versus non-responders pre-treatment. **e**, CXCL13 IHC staining of the CTCL tissue microarray. Scale bars, 100 µm. **f**, Normalized bulk *CXCR5* gene expression across patient groups. Boxes, median ± IQR; whiskers, 1.5x IQR. *P* values were calculated with a linear mixed-effect model with Bonferroni’s corrections for multiple comparisons.

**Supplementary Table 1.Cohort and tissue microarray layout.**

**Sup. Table 1a.** Cohort.

| Patient ID | Age at screening (years) | Gender | Diagnosis | Disease stage | Prior therapies | Response status | Biopsy timepoints | C02 (weeks) | Response / progression time (weeks) | EOT time (weeks) | Death time (weeks) | Follow-up time (weeks) |
| --- | --- | --- | --- | --- | --- | --- | --- | --- | --- | --- | --- | --- |
| 1 | 76.14 | Male | MF | IIIA | 3 | Responder | Pre, EOT |  |  | 104 |  | 142 |
| 2 | 60.59 | Male | MF | IIIB | 4 | Responder | Pre, EOT |  |  | 104 |  | 151 |
| 3 | 52.01 | Male | SS | IVA | 3 | Non-responder | Pre, EOT |  |  | 98 | 98 | 98 |
| 4 | 85.14 | Female | SS | IVA | 4 | Non-responder | Pre, EOT |  |  | 104 | 142 | 142 |
| 5 | 75.24 | Male | MF | IIB | 6 | Responder | Pre, EOT |  |  | 104 |  | 103 |
| 6 | 46.74 | Male | MF | IVA | 9 | Non-responder | Pre, C02, EOT | 3 |  | 25 |  | 25 |
| 7 | 46.71 | Female | MF | IIIB | 3 | Non-responder | Pre, EOT |  |  | 9 |  | 9 |
| 8 | 67.44 | Male | SS | IIIB | 1 | Non-responder | Pre, EOT |  |  | 104 | 121 | 121 |
| 9 | 77.65 | Female | SS | IVA | 6 | Responder | Pre, C02, EOT | 3 |  | 71 |  | 71 |
| 10 | 66.40 | Male | MF | IIIA | 2 | Responder | Pre |  |  | N/A | 119 | 119 |
| 11 | 72.23 | Male | SS | IVA | 3 | Responder | Pre, Response |  | 22 | 104 |  | 171 |
| 12 | 72.59 | Male | SS | IVA | 5 | Non-responder | Pre, Progression |  | 15 | 15 |  | 15 |
| 13 | 44.10 | Female | SS | IVA | 1 | Responder | Pre, C02, Response, EOT | 3 | 15 | 104 |  | 179 |
| 14 | 63.69 | Male | MF | IB | 5 | Non-responder | Pre, C02, Response, EOT | 3 | 10 | 92 | 92 | 92 |

**MF:** mycosis fungoides
**SS:** Sezary syndrome
**Pre:** biopsy obtained pre-treatment
**C02:** biopsy obtained prior to cycle 2
**Response / Progression:** biopsy obtained at point of response or progression
**EOT:** end of trial

**Sup. Table 1b.** Tissue microarray layout.

| TMA spot | Patient ID | Biopsy timepoint |
| --- | --- | --- |
| 1 | 1 | Pre_1 |
| 2 | 1 | Pre_2 |
| 3 | 1 | EOT_1 |
| 4 | 1 | EOT_2 |
| 5 | 2 | Pre_1 |
| 6 | 2 | Pre_2 |
| 7 | 2 | EOT_1 |
| 8 | 2 | EOT_2 |
| 9 | 3 | Pre_1 |
| 10 | 3 | Pre_2 |
| 11 | 3 | EOT_1 |
| 12 | 3 | EOT_2 |
| 13 | 4 | Pre_1 |
| 14 | 4 | Pre_2 |
| 15 | 4 | EOT_1 |
| 16 | 4 | EOT_2 |
| 17 | 5 | Pre_1 |
| 18 | 5 | Pre_2 |
| 19 | 5 | EOT_1 |
| 20 | 5 | EOT_2 |
| 21 | 6 | Pre_1 |
| 22 | 6 | Pre_2 |
| 23 | 6 | EOT_1 |
| 24 | 6 | EOT_2 |
| 25 | 7 | Pre_1 |
| 26 | 7 | Pre_2 |
| 27 | 7 | EOT_1 |
| 28 | 7 | EOT_2 |
| 29 | 8 | Pre_1 |
| 30 | 8 | Pre_2 |
| 31 | 8 | Pre_3 |
| 32 | 8 | EOT_1 |
| 33 | 8 | EOT_2 |
| 34 | 8 | EOT_3 |
| 35 | 9 | Pre_1 |
| 36 | 9 | Pre_2 |
| 37 | 9 | EOT_1 |
| 38 | 9 | EOT_2 |
| 39 | 10 | Pre_1 |
| 40 | 10 | Pre_2 |
| 41 | 11 | Pre_1 |
| 42 | 11 | Pre_2 |
| 43 | 11 | Resp_1 |
| 44 | 11 | Resp_2 |
| 45 | 12 | Pre_1 |
| 46 | 12 | Pre_2 |
| 47 | 12 | Resp_1 |
| 48 | 12 | Resp_2 |
| 49 | 6 | C02_1 |
| 50 | 6 | C02_2 |
| 51 | 13 | Pre_1 |
| 52 | 13 | Pre_2 |
| 53 | 13 | Resp_1 |
| 54 | 13 | Resp_2 |
| 55 | 13 | EOT_1 |
| 56 | 13 | EOT_2 |
| 57 | 14 | Pre_1 |
| 58 | 14 | Pre_2 |
| 59 | 14 | Resp_1 |
| 60 | 14 | Resp_2 |
| 61 | 14 | Resp_3 |
| 62 | 14 | EOT_1 |
| 63 | 14 | EOT_2 |
| 64 | 14 | EOT_3 |
| 65 | 14 | C02_1 |
| 66 | 14 | C02_2 |
| 67 | 9 | C02_1 |
| 68 | 9 | C02_2 |
| 69 | 13 | C02_1 |
| 70 | 13 | C02_2 |

**Pre:** pre-treatment biopsy
**C02:** biopsy obtained prior to cycle 2 (3 weeks)
**Response / Progression:** biopsy obtained at point of response or progression
**EOT:** end of trial (104 weeks)

**Supplementary Table 2.**
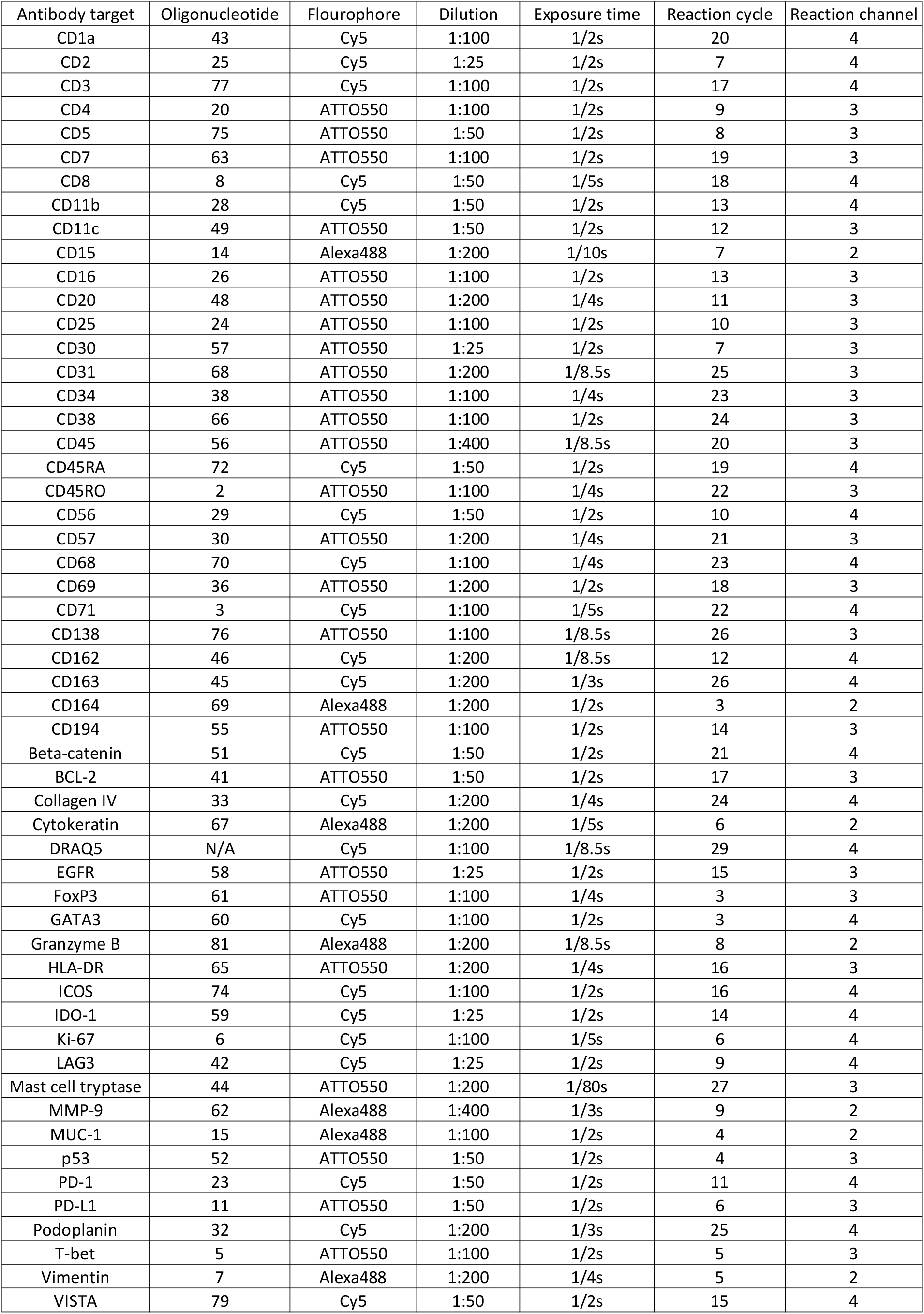
Targets.

| Antibody target | Oligonucleotide | Fluorophore | Dilution | Exposure time | Reaction cycle | Reaction channel |
| --- | --- | --- | --- | --- | --- | --- |
| CD1a | 43 | Cy5 | 1:100 | 1/2s | 20 | 4 |
| CD2 | 25 | Cy5 | 1:25 | 1/2s | 7 | 4 |
| CD3 | 77 | Cy5 | 1:100 | 1/2s | 17 | 4 |
| CD4 | 20 | ATTO550 | 1:100 | 1/2s | 9 | 3 |
| CD5 | 75 | ATTO550 | 1:50 | 1/2s | 8 | 3 |
| CD7 | 63 | ATTO550 | 1:100 | 1/2s | 19 | 3 |
| CD8 | 8 | Cy5 | 1:50 | 1/5s | 18 | 4 |
| CD11b | 28 | Cy5 | 1:50 | 1/2s | 13 | 4 |
| CD11c | 49 | ATTO550 | 1:50 | 1/2s | 12 | 3 |
| CD15 | 14 | Alexa488 | 1:200 | 1/10s | 7 | 2 |
| CD16 | 26 | ATTO550 | 1:100 | 1/2s | 13 | 3 |
| CD20 | 48 | ATTO550 | 1:200 | 1/4s | 11 | 3 |
| CD25 | 24 | ATTO550 | 1:100 | 1/2s | 10 | 3 |
| CD30 | 57 | ATTO550 | 1:25 | 1/2s | 7 | 3 |
| CD31 | 68 | ATTO550 | 1:200 | 1/8.5s | 25 | 3 |
| CD34 | 38 | ATTO550 | 1:100 | 1/4s | 23 | 3 |
| CD38 | 66 | ATTO550 | 1:100 | 1/2s | 24 | 3 |
| CD45 | 56 | ATTO550 | 1:400 | 1/8.5s | 20 | 3 |
| CD45RA | 72 | Cy5 | 1:50 | 1/2s | 19 | 4 |
| CD45RO | 2 | ATTO550 | 1:100 | 1/4s | 22 | 3 |
| CD56 | 29 | Cy5 | 1:50 | 1/2s | 10 | 4 |
| CD57 | 30 | ATTO550 | 1:200 | 1/4s | 21 | 3 |
| CD68 | 70 | Cy5 | 1:100 | 1/4s | 23 | 4 |
| CD69 | 36 | ATTO550 | 1:200 | 1/2s | 18 | 3 |
| CD71 | 3 | Cy5 | 1:100 | 1/5s | 22 | 4 |
| CD138 | 76 | ATTO550 | 1:100 | 1/8.5s | 26 | 3 |
| CD162 | 46 | Cy5 | 1:200 | 1/8.5s | 12 | 4 |
| CD163 | 45 | Cy5 | 1:200 | 1/3s | 26 | 4 |
| CD164 | 69 | Alexa488 | 1:200 | 1/2s | 3 | 2 |
| CD194 | 55 | ATTO550 | 1:100 | 1/2s | 14 | 3 |
| Beta-catenin | 51 | Cy5 | 1:50 | 1/2s | 21 | 4 |
| BCL-2 | 41 | ATTO550 | 1:50 | 1/2s | 17 | 3 |
| Collagen IV | 33 | Cy5 | 1:200 | 1/4s | 24 | 4 |
| Cytokeratin | 67 | Alexa488 | 1:200 | 1/5s | 6 | 2 |
| DRAQ5 | N/A | Cy5 | 1:100 | 1/8.5s | 29 | 4 |
| EGFR | 58 | ATTO550 | 1:25 | 1/2s | 15 | 3 |
| FoxP3 | 61 | ATTO550 | 1:100 | 1/4s | 3 | 3 |
| GATA3 | 60 | Cy5 | 1:100 | 1/2s | 3 | 4 |
| Granzyme B | 81 | Alexa488 | 1:200 | 1/8.5s | 8 | 2 |
| HLA-DR | 65 | ATTO550 | 1:200 | 1/4s | 16 | 3 |
| ICOS | 74 | Cy5 | 1:100 | 1/2s | 16 | 4 |
| IDO-1 | 59 | Cy5 | 1:25 | 1/2s | 14 | 4 |
| Ki-67 | 6 | Cy5 | 1:100 | 1/5s | 6 | 4 |
| LAG3 | 42 | Cy5 | 1:25 | 1/2s | 9 | 4 |
| Mast cell tryptase | 44 | ATTO550 | 1:200 | 1/80s | 27 | 3 |
| MMP-9 | 62 | Alexa488 | 1:400 | 1/3s | 9 | 2 |
| MUC-1 | 15 | Alexa488 | 1:100 | 1/2s | 4 | 2 |
| p53 | 52 | ATTO550 | 1:50 | 1/2s | 4 | 3 |
| PD-1 | 23 | Cy5 | 1:50 | 1/2s | 11 | 4 |
| PD-L1 | 11 | ATTO550 | 1:50 | 1/2s | 6 | 3 |
| Podoplanin | 32 | Cy5 | 1:200 | 1/3s | 25 | 4 |
| T-bet | 5 | ATTO550 | 1:100 | 1/2s | 5 | 3 |
| Vimentin | 7 | Alexa488 | 1:200 | 1/4s | 5 | 2 |
| VISTA | 79 | Cy5 | 1:50 | 1/2s | 15 | 4 |

**Supplementary Table 3.Genes lists; related to Figures 2e-h, 4f-h.**

**Sup. Table 3a.** Interferon gamma gene score^39^.

| Gene | Coefficient |
| --- | --- |
| IFNG | -0.30685 |
| HLA-DRA | -0.35484 |
| IDO1 | -0.36499 |
| STAT1 | -0.45212 |
| CXCL10 | -0.46207 |
| CXCL9 | -0.47832 |

**Sup. Table 3b.**
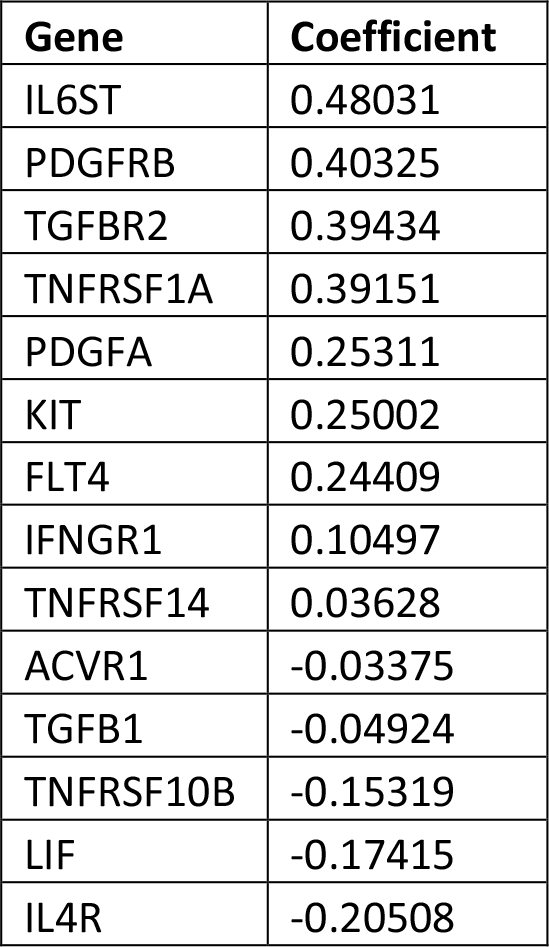
Transforming growth factor beta gene score^40^.

| Gene | Coefficient |
| --- | --- |
| IL6ST | 0.48031 |
| PDGFRB | 0.40325 |
| TGFBR2 | 0.39434 |
| TNFRSF1A | 0.39151 |
| PDGFA | 0.25311 |
| KIT | 0.25002 |
| FLT4 | 0.24409 |
| IFNGR1 | 0.10497 |
| TNFRSF14 | 0.03628 |
| ACVR1 | -0.03375 |
| TGFB1 | -0.04924 |
| TNFRSF10B | -0.15319 |
| LIF | -0.17415 |
| IL4R | -0.20508 |

**Sup. Table 3c.** Immune activation gene score.

| Gene | Coefficient |
| --- | --- |
| CXCL9 | 0.28056 |
| CCL5 | 0.27507 |
| GZMH | 0.27229 |
| EOMES | 0.26916 |
| GZMK | 0.26379 |
| CD27 | 0.25655 |
| TNFRSF9 | 0.25375 |
| IL2RG | 0.24956 |
| PRF1 | 0.24024 |
| FASLG | 0.23683 |
| IL2RA | 0.22805 |
| IFNG | 0.21895 |
| ICOS | 0.20233 |
| CD40 | 0.18924 |
| CD40LG | 0.18919 |
| IL17RA | 0.14444 |
| IL12B | 0.13478 |
| TNFSF9 | 0.12999 |
| CD28 | 0.12862 |
| CX3CR1 | 0.11662 |
| TNFRSF4 | 0.11086 |
| GZMB | 0.07612 |
| CCR7 | 0.06002 |
| IL23A | 0.05400 |

**Sup. Table 3d.** Immunosuppression gene score.

| Gene | Coefficient |
| --- | --- |
| TGFB1 | 0.31255 |
| ENTPD1 | 0.30707 |
| IL24 | 0.30658 |
| CXCL1 | 0.30076 |
| IL6 | 0.29989 |
| TGFBR1 | 0.27375 |
| IL10 | 0.24295 |
| HAVCR2 | 0.20469 |
| PDGFRA | 0.20016 |
| HLA-G | 0.18822 |
| TGFBRAP1 | 0.18282 |
| PVT1 | 0.16990 |
| PDGFRB | 0.16723 |
| LINC00473 | 0.16219 |
| CXCL3 | 0.16065 |
| TIGIT | 0.15326 |
| LIF | 0.14962 |
| CXCL12 | 0.12772 |
| CD274 | 0.12391 |
| TIAF1 | 0.11831 |
| LAG3 | 0.10954 |
| DNM3OS | 0.02112 |
| NIFK-AS1 | -0.00826 |
| IL4 | -0.04139 |
| GNAS-AS1 | -0.05070 |
| HOTAIR | -0.06027 |
| PDCD1LG2 | -0.08477 |
| GAS5 | -0.14413 |

**Sup. Table 3e.** Cytotoxicity gene score^49,50^.

| Gene | Coefficient |
| --- | --- |
| NKG7 | 0.39045 |
| PRF1 | 0.35018 |
| TNF | 0.34146 |
| GZMH | 0.32965 |
| TBX21 | 0.31663 |
| GZMK | 0.31526 |
| GZMM | 0.29080 |
| ID2 | 0.26046 |
| IFNG | 0.25047 |
| GNLY | 0.22788 |
| GZMB | 0.18149 |

**Sup. Table 3f.** Tumor aggressiveness^51,52^.

| Gene | Coefficient |
| --- | --- |
| IL2RB | 0.30377 |
| BATF | 0.29598 |
| IL21R | 0.28415 |
| CCND2 | 0.26500 |
| RGS16 | 0.25940 |
| MYO7A | 0.25910 |
| SLA | 0.25436 |
| ANP32E | 0.23795 |
| MTHFD2 | 0.21538 |
| IL10 | 0.21012 |
| TRIB2 | 0.20521 |
| RARRES3 | 0.19264 |
| GNLY | 0.19238 |
| DUSP5 | 0.18767 |
| C1GALT1 | 0.15656 |
| BCL2 | 0.15272 |
| EHD1 | 0.13197 |
| HSPD1 | 0.13011 |
| DAD1 | 0.12384 |
| MMP12 | 0.08265 |
| IL26 | 0.07073 |
| IL22 | 0.06240 |
| P4HB | -0.05212 |
| TCN1 | -0.07340 |
| SFTPD | -0.12905 |
| TGFBR3 | -0.18280 |

**Supplementary Table 4.CIBERSORTx marker genes.**

**Sup. Table 4a.** Markers of CIBERSORTx cell-types.

| Cell-types | Marker genes | CIBERSORTx cell-types |
| --- | --- | --- |
| B cells & plasma cells | *MS4A1, LTB, CD79A, CD79B* | B and plasma cells |
| T cells | *CD3E, CD3D, CD274* | CD4 T, CD8 T, Tregs, tumor cells |
| Macrophages & dendritic cells | *AIF1* | Macrophages, dendritic cells |
| Keratinocytes | *KRT1, KRT14* | Epithelium |
| Vasculature | *CD43, PECAM1, CLDN5* | Vasculature |
| Fibroblasts | *COL1A1, SFRP2* | Stroma |
| Pericytes | *RGS5, ACTA2* |  |

**Sup. Table 4b.**
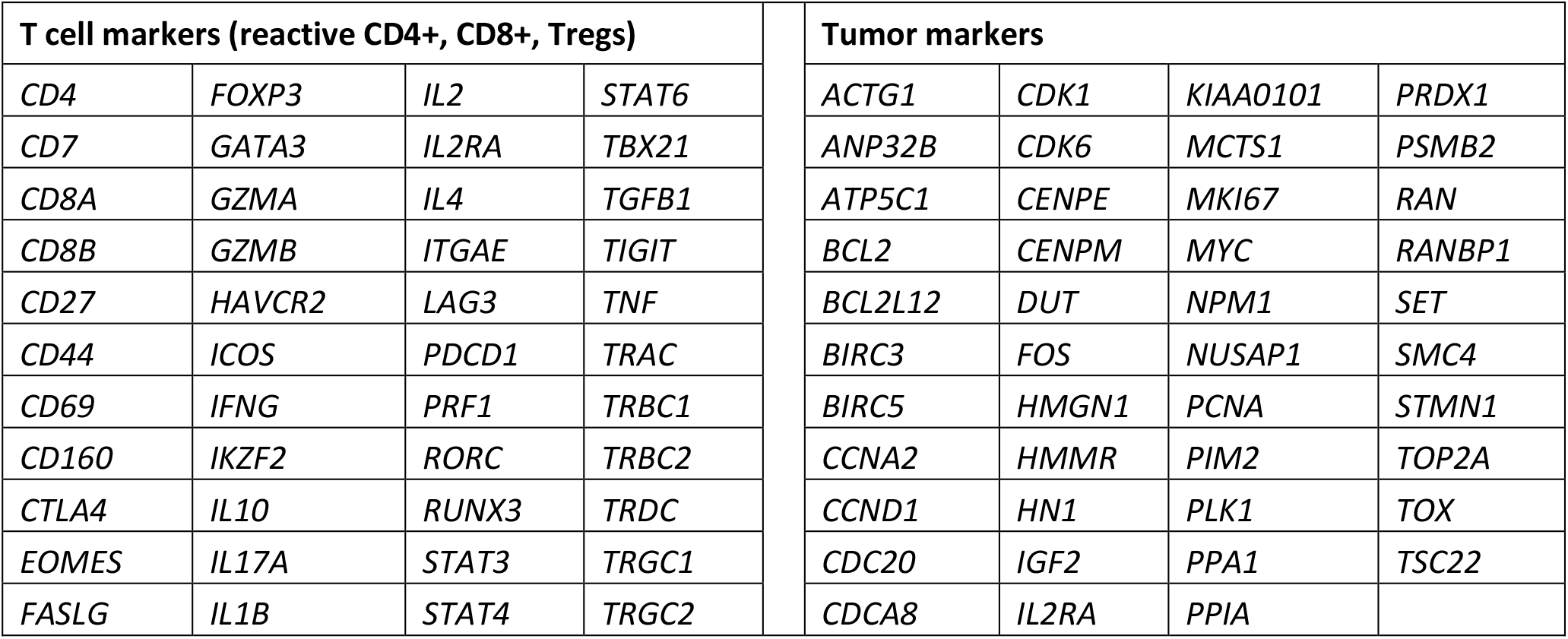
T cell and tumor cell markers for CIBERSORTx.

